# Neutrophil proteomics identifies temporal changes and hallmarks of delayed recovery in COVID19

**DOI:** 10.1101/2022.08.21.22279031

**Authors:** Merete B Long, Andrew JM Howden, Holly R Keir, Christina M Rollings, Yan Hui Giam, Thomas Pembridge, Lilia Delgado, Hani Abo-Leyah, Amy F Lloyd, Gabriel Sollberger, Rebecca Hull, Amy Gilmour, Chloe Hughes, Benjamin JM New, Diane Cassidy, Amelia Shoemark, Hollian Richardson, Angus I Lamond, Doreen A Cantrell, James D Chalmers, Alejandro J Brenes

**Affiliations:** Division of Molecular and Clinical Medicine, University of Dundee, Ninewells Hospital and Medical School, Dundee, Scotland, UK; Division of Cell Signalling and Immunology, School of Life Sciences, University of Dundee, Dundee, Scotland, UK; Centre for Gene Regulation and Expression, School of Life Sciences, University of Dundee, Dundee, Scotland, UK; Max Planck Institute for Infection Biology, Berlin, Germany; Department of Infection, Immunity and Cardiovascular Disease, University of Sheffield, UK

**Keywords:** Neutrophil, proteomics, Interferon, long covid

## Abstract

**Rationale:** Neutrophils are important in the pathophysiology of COVID19 but the molecular changes contributing to altered neutrophil phenotypes following SARS-CoV-2 infection are not fully understood.

**Objectives:** To use quantitative mass spectrometry-based proteomics to explore neutrophil phenotypes following acute SARS-CoV-2 infection and during recovery.

**Methods:** Prospective observational study of hospitalised patients with PCR-confirmed SARS-CoV-2 infection (May 2020-December 2020). Patients were enrolled within 96 hours of admission, with longitudinal sampling up to 29 days. Control groups comprised non-COVID19 acute lower respiratory tract infection (LRTI) and age-matched non-infected controls. Neutrophils isolated from peripheral blood were processed for mass spectrometry. COVID19 severity and recovery were defined using the WHO ordinal scale.

**Measurements and Main Results:** 84 COVID19 patients were included and compared to 91 LRTI patients and 42 controls. 5,800 neutrophil proteins were identified and 1,748 proteins were significantly different (q-value<0.05) in neutrophils from COVID19 patients compared to those of non-infected controls, including a robust interferon response at baseline, which was lost in severe patients one week after enrolment. Neutrophil changes associated with COVID19 disease severity and prolonged illness were characterized and candidate targets for modulation of neutrophil function were identified. Delayed recovery from COVID19 was associated with changes in metabolic and signalling proteins, complement, chemokine and leukotriene receptors, integrins and inhibitory receptors.

**Conclusions:** SARS-CoV-2 infection results in the sustained presence of recirculating neutrophils with distinct metabolic profiles and altered capacities to respond to migratory signals and cues from other immune cells, pathogens or cytokines.

**Scientific Knowledge on the Subject:** Inflammation is the primary driver of morbidity and mortality in severe COVID19. Type I interferon responses, T-cell exhaustion, cytokine storm, emergency myelopoiesis, myeloid compartment dysregulation and procoagulant pathway activation are well established contributors to COVID19 disease severity. Neutrophils play an important role in COVID19, with elevated neutrophil-to-lymphocyte ratios and the emergence of a circulating immature neutrophil population in individuals with severe symptoms. Neutrophil infiltration in the lungs coupled with the release of neutrophil extracellular traps has also been reported in severe and fatal COVID19. The aim of this study was to quantitatively map the proteomes of peripheral blood neutrophils from a cohort of hospitalised COVID19 patients to understand how SARS-CoV-2 infection changes neutrophil phenotypes and functional capacity.

**What this study adds to the field:** High-resolution mass spectrometry was used to characterise the proteomes of peripheral blood neutrophils from >200 individuals at different stages of disease. This work has comprehensively mapped neutrophil molecular changes associated with mild and severe COVID19 and identified significant quantitative changes in more than 1700 proteins in neutrophils from patients hospitalised with COVID19 versus patients with non-COVID19 acute respiratory infections. The study identifies neutrophil protein signatures associated with COVID19 disease severity. The data also show that alterations in neutrophil proteomes can persist in fully recovered patients and identify distinct neutrophil proteomes in recovered versus non recovered patients. Our study provides novel insights into neutrophil responses during acute COVID19 and reveals that altered neutrophil phenotypes persist in convalescent COVID19 patients.

## Introduction

Coronavirus disease 2019 (COVID19) caused by SARS-CoV-2 infection has a diverse spectrum of presentations; from asymptomatic or pre-symptomatic, to mild illness with flu-like symptoms, moderate illness demonstrating lower respiratory disease and severe or critical illness involving development of pneumonia and acute respiratory distress syndrome (ARDS)^(1-3)^. The immune response plays a key role in determining the outcome of SARS-CoV-2 infection, with an excessive inflammatory response accounting for the majority of morbidity and mortality. Major immune events following COVID19 include generation of a type I interferon response (IFN-I)^(4, 5)^, reduced numbers of T cells^(6-8)^, cytokine storm^(6, 7, 9, 10)^, emergency myelopoiesis and myeloid compartment dysregulation^(4, 8, 11, 12)^, as well as procoagulant pathway activation^(10, 13)^. A hallmark of severe disease is T-cell lymphopenia in combination with increased blood neutrophil counts^(2, 14-16)^. Neutrophils are key effector cells of the innate immune response that play a role in responses to pathogens and tumours^(17, 18)^ and in severe COVID19 there is evidence of emergency myelopoiesis and the appearance of circulating immature and dysfunctional neutrophils in peripheral blood^(11, 19-22)^. There is also a signature of neutrophil activation in patients with severe COVID19 that predicts disease trajectory^(23, 24)^, as well as evidence of increased NETosis^(25)^. Neutrophil populations following SARS-CoV-2 infection have been characterized by combinations of high-resolution mass cytometry^(22, 26, 27)^, flow cytometry^(21)^ and by single cell RNA sequencing^(4, 28-30)^. However, it is recognised that in-depth quantitative analysis of cellular proteomes can provide insights unobtainable from transcriptomes, particularly relating to neutrophil biology^(31)^. Proteomic analysis of neutrophils has thus identified a type 1 interferon and prothrombotic hyperinflammatory signature in neutrophils isolated from COVID19 infected individuals with acute respiratory distress syndrome (ARDS)^(32)^. However, there has been no systematic analysis of neutrophil proteomes in patients with COVID19 of differing severity and no exploration of the persistence of changes in neutrophil phenotypes following SARS-CoV-2 infection. Accordingly, in the current study mass spectrometry was used to map the protein signatures of peripheral blood neutrophils from a cohort of patients who were admitted to hospital with COVID19 from May 2020 to December 2020, sampled up to 29 days following hospitalization, with matched control populations. This extensive patient cohort and sampling strategy has provided an in-depth analysis of the neutrophil proteomes in health and disease and provided novel insights into changes in neutrophils during acute SARS-CoV-2 infection and during the disease recovery phase.

## Methods

The PREDICT-COVID19 study was a prospective observational case-control study conducted at Ninewells Hospital, Dundee, UK. Patients with suspected or confirmed COVID19 were enrolled within 96 hours of hospital admission with SARS-CoV-2 infection confirmed by RT-PCR performed on combined oropharyngeal and nasopharyngeal swabs. The study was approved by the East of Scotland Research Ethics Committee (20/ES/0055) and written informed consent was provided by all participants. Two control populations were included; (i) patients presenting with community-acquired lower respiratory tract infections (LRTI) not due to SARS-CoV-2 infection and (ii) age and sex matched, non-infected controls.

Blood sampling was performed at enrolment (study day 1), and for the COVID19 cohort additional blood sampling was performed on day 7, 15 and 29 while in hospital. The timing of blood sampling was standardised (between 0900-1100h each day). A subset of participants who had been discharged returned as outpatients for sampling at day 29. Full inclusion and exclusion criteria are shown online. Inclusion criteria for controls were age ≥16 years, absence of an infection-related diagnosis, judged as clinically stable by the investigator and able to give informed consent. Exclusion criteria were known or past SARS-CoV-2 infection in the past 3 months, known contact with a COVID19 positive case in the preceding 14 days, any current infection, and any contraindication to venepuncture or participation in the study as judged by the investigator.

### Clinical variables

Baseline severity was classified according to the WHO scale as WHO3= Hospitalized, not requiring supplementary oxygen, WHO4= Hospitalized, requiring oxygen through facemask or nasal prongs, WHO5/6= requiring high flow nasal oxygen, continuous positive airway pressure or invasive mechanical ventilation. Patients were categorised as either recovered (WHO=1) or not recovered (WHO2-3) at day 29, as determined on the basis of symptoms reported at follow-up, with patients still hospitalized categorized as non-recovered.

### Neutrophil and PBMC isolation and sample preparation for LC-MS

Within two hours of venepuncture, neutrophils were isolated using the EasySep(tm) Direct Human Neutrophil Isolation Kit (STEMCELL Technologies #19666) utilising negative immunomagnetic selection and peripheral blood mononuclear cell (PBMCs) isolated by density-gradient separation using Lymphoprep™ and SepMate™ columns. Isolated cells were pelleted and stored at -80°C until analysis. To minimise batch effects of lysis buffers and conditions, stored cell pellets (5×10^6^ cells) were lysed in batches of 50–100 samples.

### Proteomic sample preparation and mass spectrometry analysis

Neutrophil and PBMC pellets were processed for mass spectrometry as described^(32)^. For each sample, 1.5 μg of peptide was analysed on a Q Exactive HF-X (Thermo Scientific) mass spectrometer coupled with a Dionex Ultimate 3000 RS (Thermo Scientific). The mass spectrometer was operated in data-independent acquisition mode (DIA), and the raw MS data were processed using Spectronaut^(33)^. Full details are described in the supplementary methods.

### Statistical methods

The differential expression analyses were performed in R (v. 4.0.3) and the global p-values and fold changes were calculated via the Bioconductor package Limma^(34)^ (v 3.46.0). The q-value was calculated with the Bioconductor package qvalue (v 2.22.0). Differences in global protein levels were considered significant when the q-value≤0.05. P-values for protein families and the PBMC proteins were calculated in R using Welches T-test, with differences considered significant when the p-value≤0.05. Overrepresentation analyses were performed using webgestalt^(35)^, full details are described in the supplementary methods.

## Results

217 patients were enrolled between May 2020 and December 2020. The cohort consisted of 84 individuals with confirmed COVID-19, 91 with acute LRTI and 42 non-infected controls (Table 1). The COVID19 and control populations were well matched at the baseline (day 1), with slightly more females in the uninfected control population.

**Table 1.**
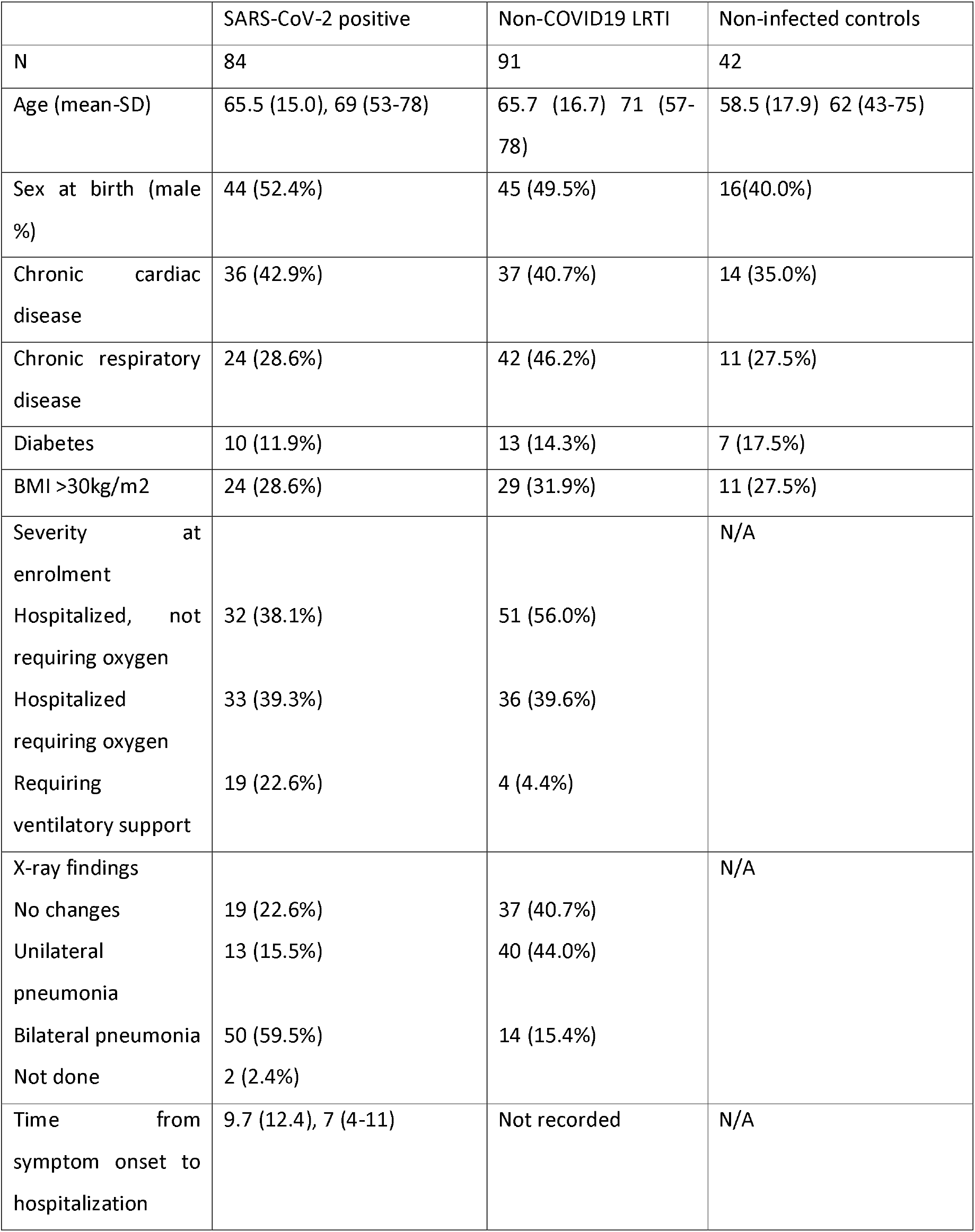
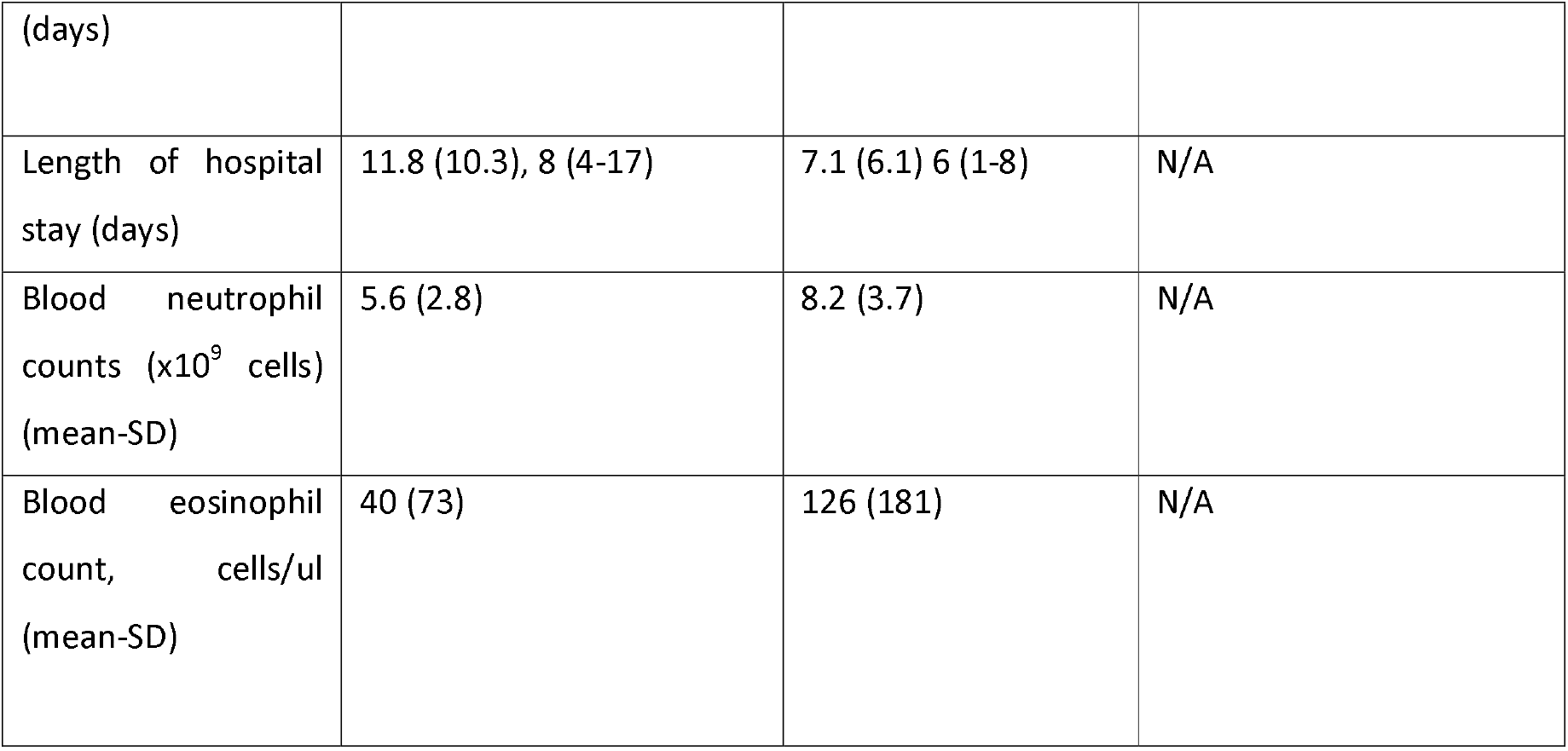
Patient characteristics at enrolment At baseline 32 (38.1%) patients with COVID19 did not require supplementary oxygen, 33 (39.3%) required supplementary oxygen but did not require ventilatory support and 19 (22.6%) required mechanical ventilation or non-invasive ventilatory support. 46 patients received 6mg dexamethasone daily (54.8%).

|  | SARS-CoV-2 positive | Non-COVID19 LRTI | Non-infected controls |
| --- | --- | --- | --- |
| N | 84 | 91 | 42 |
| Age (mean-SD) | 65.5 (15.0), 69 (53-78) | 65.7 (16.7) 71 (57-78) | 58.5 (17.9) 62 (43-75) |
| Sex at birth (male %) | 44 (52.4%) | 45 (49.5%) | 16(40.0%) |
| Chronic cardiac disease | 36 (42.9%) | 37 (40.7%) | 14 (35.0%) |
| Chronic respiratory disease | 24 (28.6%) | 42 (46.2%) | 11 (27.5%) |
| Diabetes | 10 (11.9%) | 13 (14.3%) | 7 (17.5%) |
| BMI >30kg/m2 | 24 (28.6%) | 29 (31.9%) | 11 (27.5%) |
| Severity at enrolment |  |  | N/A |
| Hospitalized, not requiring oxygen | 32 (38.1%) | 51 (56.0%) |  |
| Hospitalized requiring oxygen | 33 (39.3%) | 36 (39.6%) |  |
| Requiring ventilatory support | 19 (22.6%) | 4 (4.4%) |  |
| X-ray findings |  |  | N/A |
| No changes | 19 (22.6%) | 37 (40.7%) |  |
| Unilateral pneumonia | 13 (15.5%) | 40 (44.0%) |  |
| Bilateral pneumonia | 50 (59.5%) | 14 (15.4%) |  |
| Not done | 2 (2.4%) |  |  |
| Time from symptom onset to hospitalization | 9.7 (12.4), 7 (4-11) | Not recorded | N/A |
| (days) |  |  |  |
| Length of hospital stay (days) | 11.8 (10.3), 8 (4-17) | 7.1 (6.1) 6 (1-8) | N/A |
| Blood neutrophil counts (x10<sup>9</sup> cells) (mean-SD) | 5.6 (2.8) | 8.2 (3.7) | N/A |
| Blood eosinophil count, cells/ul (mean-SD) | 40 (73) | 126 (181) | N/A |

### The protein landscape of neutrophils from hospitalised COVID19 patients

Peripheral blood neutrophil proteomes from patients with COVID19, non-COVID19 LRTI and control groups were analysed by mass spectrometry (Fig 1a). Over 5,800 unique proteins were identified with a median of 4,923 proteins/sample (Fig 1b) allowing both quantitative and qualitative comparisons of neutrophil proteomes. There were no significant differences in the total protein content of neutrophils from control, LRTI or COVID19 patients (Fig. 1c) but there were differences in their protein landscape composition. There were 300 proteins significantly increased and 123 proteins significantly decreased in abundance in neutrophils from LTRI patients compared to controls (Fig. 1d; S.Table 1). These changes include decreases in abundance of CD10 (MME) and components of azurophilic granules including elastase and myeloperoxidase. Proteins increased in abundance control vesicular trafficking and mRNA processing (S.Table 2). When the proteomes of neutrophils from COVID19 patients were compared to controls, changes in expression of 1,748 proteins were detected (Fig. 1e; S. Table 3). There was increased expression of 1,008 proteins including a striking signature of interferon-regulated proteins as well as changes in proteins controlling metabolic processes including glycolysis and fatty acid metabolism. Proteins decreased in expression in neutrophils from COVID19 patients included CD10, components of endosomal sorting complexes and key enzymes controlling glycogenolysis.

**Figure 1.**
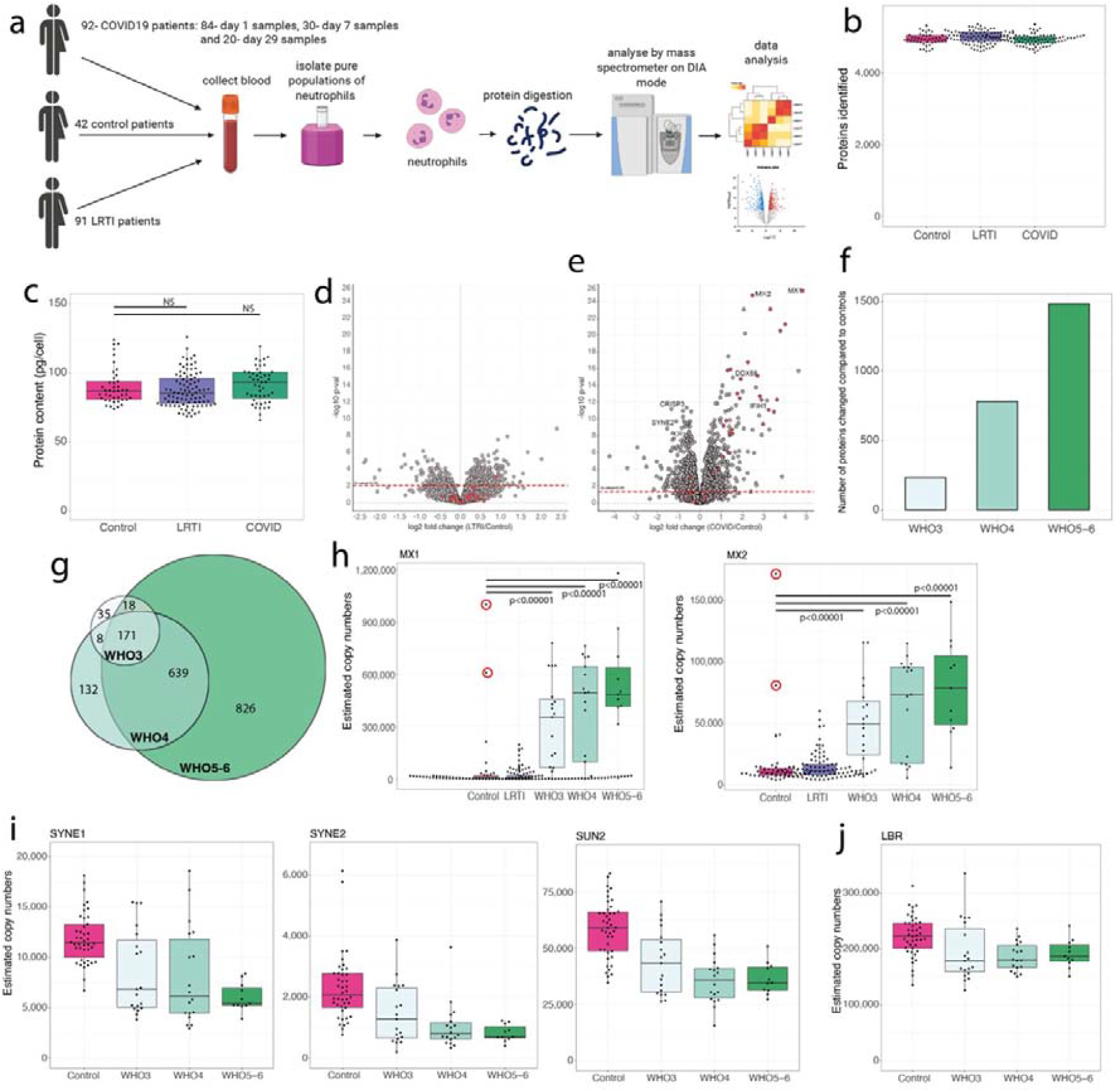
Core COVID19 neutrophil proteomic signature: (a) Sample collection and processing workflow. **(b)** Number of proteins identified across all samples for control, LRTI and COVID19. **(c)** Estimated protein content for all samples for control, LRTI and COVID19. **(d)** Volcano plot showing the fold change and p-value comparing the neutrophil proteomes of LRTI patients to the controls. IFN-induced proteins are coloured in red. The red dotted line represents Q-value=0.05. **(e)** Neutrophil proteomes of COVID19 patients compared to the controls. IFN induced proteins are coloured in red. The red dotted line represents Q-value=0.05. **(f)** Number of proteins that are significantly changed in abundance when comparing the neutrophil proteomes of WHO3 (moderate), WHO4 (severe) and WHO5-6 (critically severe) COVID19 patients to the controls. **(g)** Overlap of significantly altered proteins across the stratified COVID19 patient cohorts. **(h)** Estimated protein copy numbers for MX1 and MX2 across control, LRTI, WHO3, WHO4 and WHO5-6 COVID19 patients. Patients circled in red were pre-symptomatic and later tested SARS-CoV-2 positive. Estimated protein copy numbers for **(i)** SYNE1, SYNE2 and SUN2 and **(j)** laminin B receptor (LBR) across control, WHO3, WHO4 and WHO5-6 patients. For all boxplots the whiskers extend from the hinge to the largest and smaller values no further than 1.5 x interquartile range.

In further analysis, the COVID19 neutrophil proteomes were stratified based on the WHO severity scale upon hospital admission. This stratification revealed that the number of proteins changing increased with disease severity: 221 proteins were significantly changed in neutrophils from WHO3 patients (S.Table 4), 779 in WHO4 (S.Table 5) and 1,483 in WHO5-6 patient neutrophils (Fig. 1f; S.Table 6). There was a core signature of 171 neutrophil proteins changed across all COVID19 groups (Fig. 1g; S.Table 7). This included 101 proteins that were significantly increased in abundance which largely represented an IFN-I response present in the majority of neutrophils from WHO3, WHO4 and WHO5-6 patients (Fig. 1h; S.Fig. 1). Furthermore, this IFN response clearly distinguished the proteomes of COVID19 patients from those of the LTRI and control cohorts, even identifying two pre-symptomatic COVID19 patients originally enrolled in the control group (Fig. 1h). The core signature also included 70 proteins that were decreased in all stratified COVID19 patient cohorts. These had diverse functions and included proteins linked to the structure and rigidity of the nucleus, e.g., SYNE1, SYNE2 and SUN2 (Fig. 1i) and the laminin b receptor (LBR; Fig. 1j).

### Neutrophil phenotypes linked to COVID19 disease severity

One well established consequence of COVID19 is emergency myelopoiesis and the release of immature neutrophils into circulation^(16)^. In this respect, the current proteomic analysis revealed reduced expression of CD10 (Fig. 2a) and cytosolic components of the NADPH oxidase complex (Fig. 2b) along with increased expression of the proliferation marker PCNA in neutrophils from WHO4 and WHO5-6 patients (Fig. 2c), characteristic of immature neutrophils^(31, 36)^. Immature neutrophils are also known for their reduced density, thus can be found in the PBMC layer upon density-gradient separation^(37, 38)^. We used mass spectrometry to analyse PBMC proteomes derived from these patients. Many previous studies have shown PBMC abnormalities in patients with COVID19^(11, 12, 28, 39-43)^. Accordingly, the PBMC proteomic data highlight how COVID19 causes a disturbance of the PBMC composition with evidence for decreased levels of T cells in WHO5-6 patients and increases in MZB1^+^ cells in all COVID19 patients (S.Fig. 2). The PBMC data also showed increased expression of key neutrophil proteins such as neutrophil elastase in the PBMCs of WHO5-6 patients (Fig. 2d). The increased presence of more low-density neutrophils is consistent with an increased frequency of immature neutrophils.

**Figure 2.**
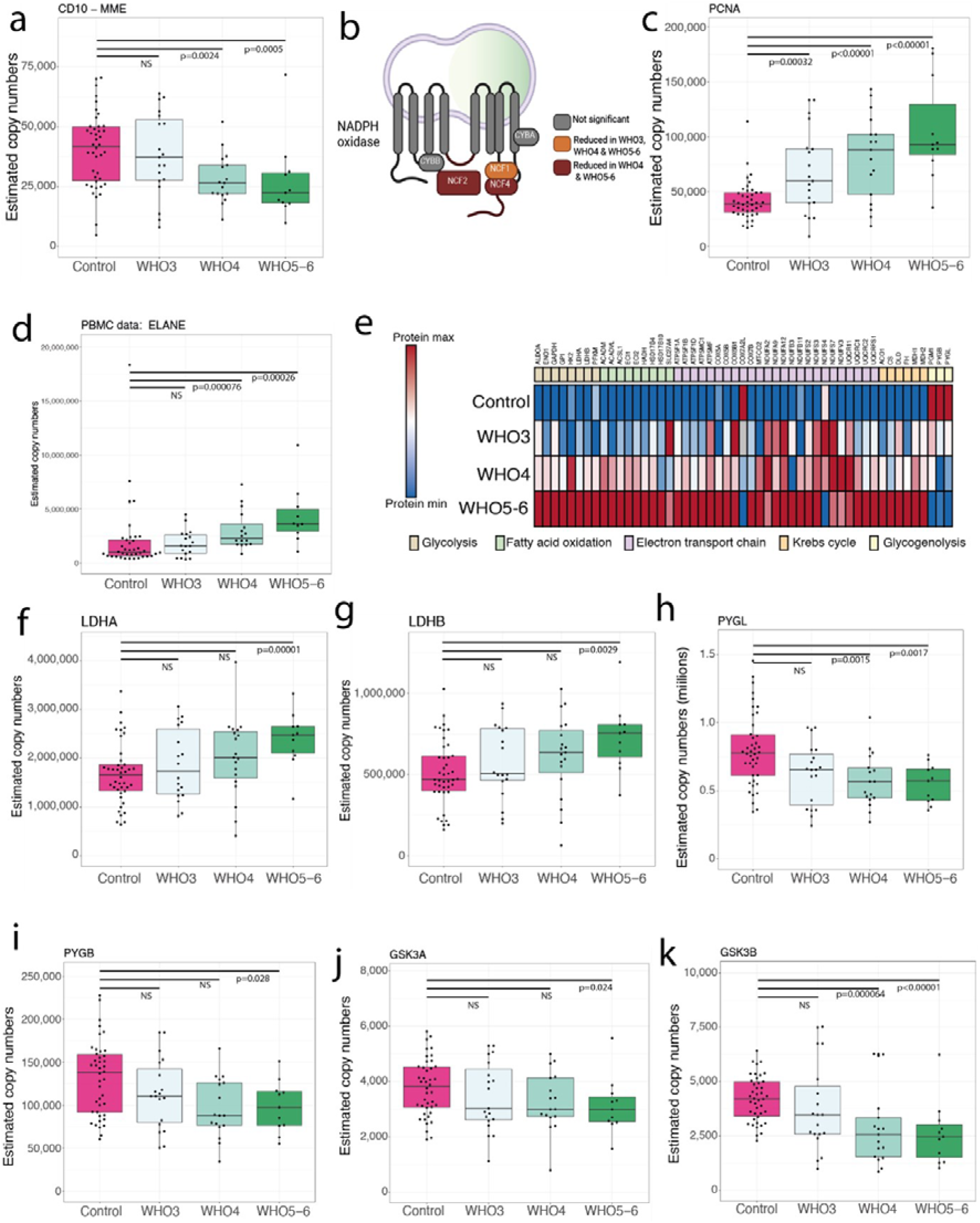
Immature neutrophils and metabolic changes in COVID19: **(a)** Estimated protein copy numbers for CD10 (MME) across control, WHO3 (moderate), WHO4 (severe) and WHO5-6 (critically ill) COVID19 patients. **(b)** NAPDH oxidase and proteins that are significantly changed in abundance compared to the control group. **(c)** Estimated protein copy numbers for PCNA across control, WHO3, WHO4 and WHO5-6 patients. **(d)** Estimated protein copy numbers for ELANE in the PBMC proteomes across control, WHO3, WHO4 and WHO5-6 patients. **(e)** Metabolic proteins that were significantly changed in abundance in WHO5-6 COVID19 patients. The data is normalised to the maximum value of each protein. Estimated protein copy numbers for **(f)** LDHA, **(g)** LDHB, **(h)** PYGL, **(i)** PYGB, **(j)** GSK3A, **(k)** GSK3B in the neutrophil proteomes across control, WHO3, WHO4 and WHO5-6 patients. For all boxplots the whiskers extend from the hinge to the largest and smaller values no further than 1.5 x interquartile range.

We also observed changes in the metabolic profile of neutrophils from WHO5-6 patients linked to neutrophil maturation stage and the effects of hypoxia^(44)^. The neutrophil proteomes from WHO5-6 patients had increased expression of enzymes involved in fatty acid oxidation, electron transport chain and the Krebs cycle (Fig. 2e), all consistent with an increased presence of immature neutrophils^(45, 46)^. However, neutrophil proteomes from WHO5-6 patients also displayed significantly higher abundance of lactate dehydrogenase A (LDHA; Fig. 2f)) and B (LDHB; Fig. 2g), vital glycolytic enzymes that convert pyruvate into lactate, which have been reported to be increased in hypoxia^(47)^. Metabolic profiling also highlighted changes related to glycogen synthesis and breakdown, where rate limiting enzymes of glycogenolysis^(48)^, glycogen phosphorylases PYGL (Fig. 2h) and PYGB (Fig. 2i) and the negative regulators of glycogen synthesis the glycogen synthase kinases^(49)^ (Fig. 2j-k) were significantly decreased in abundance WHO5-6 patients compared to controls.

Neutrophils from COVID19 patients additionally displayed changes in surface receptors (Fig. 3a), with decreased expression of migratory receptors C5AR1 and CXCR2, and increased abundance of CD64 (FCGR1A), a known neutrophil activation marker^(50, 51)^ across all patient groups (Fig. 3a). Some of the changes in these receptors associated with disease severity. There were thus increases in abundance in the proteomes of WHO5-6 patients of IL1R2, a decoy receptor for IL1 (Fig. 3b), the pattern recognition receptor TLR2 (Fig. 3c) and V-domain Ig suppressor of T cell activation (VISTA, VSIR) (Fig 3d), a negative immune checkpoint regulator. The data also highlighted reduced abundance of MHC-class II proteins that also associated with disease severity (Fig. 3e).

**Figure 3.**
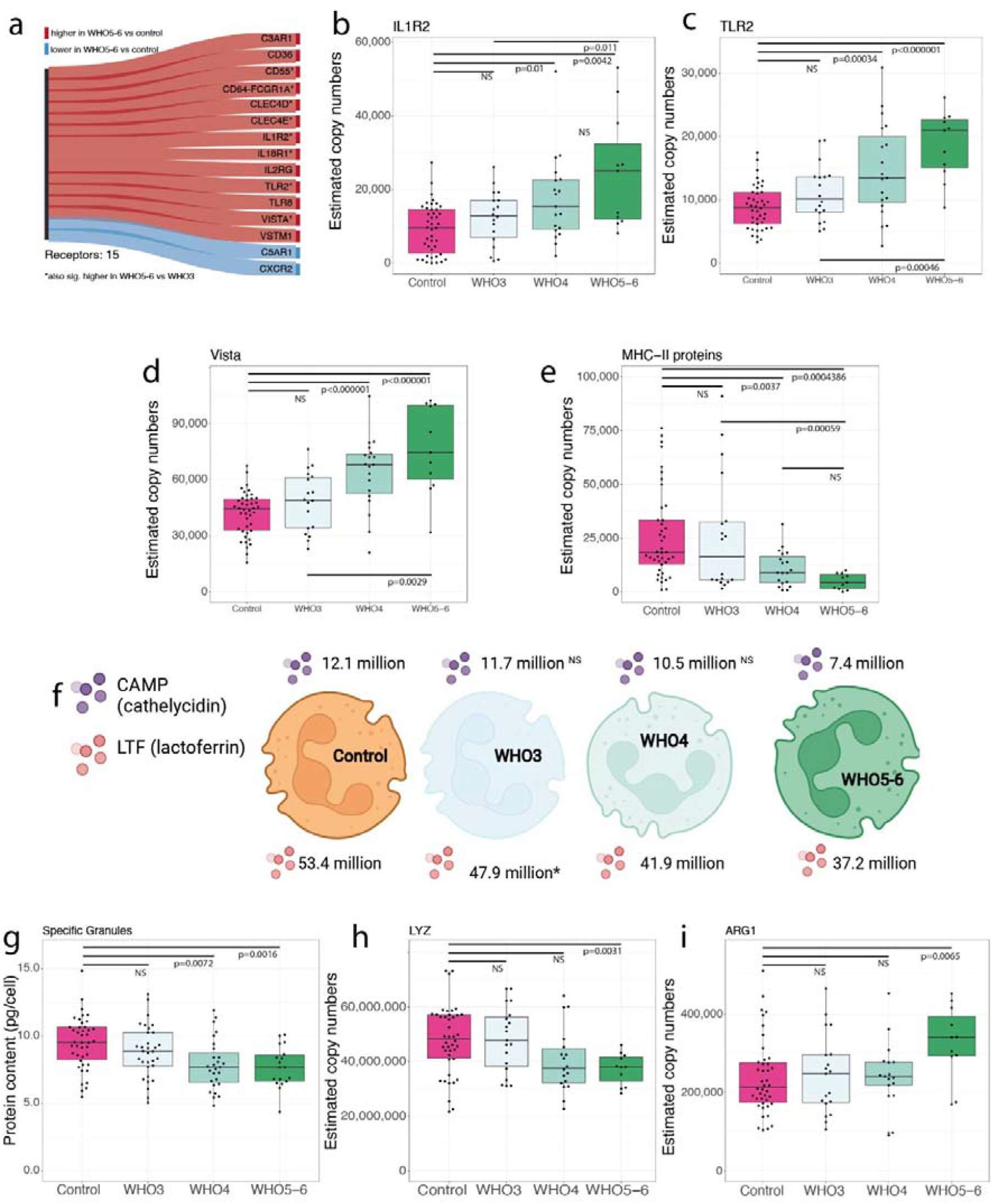
Markers of severity in the neutrophil proteomes of COVID19 patients: **(a)** Immunomodulatory receptors significantly altered in the neutrophil proteomes WHO5-6 compared to controls. Proteins in red are significantly increased in abundance in the neutrophil proteomes of WHO5-6, proteins in blue were significantly decreased in abundance. Estimated protein copy numbers for **(b)** IL1R2, **(c)** TLR2, **(d)** Vista (VSIR) **(e)** all MHC-II class proteins across control, WHO3 (moderate), WHO4 (severe) and WHO5-6 (critically ill) COVID19 patients. **(f)** Estimated median copy numbers of LTF and CAMP across control, WHO3, WHO4 and WHO5-6 patients. **(g)** Estimated protein content for proteins primarily contained within Specific Granules across control, WHO3, WHO4 and WHO5-6 patients. Estimated protein copy numbers for **(h)** LYZ and **(i)** ARG1 across control, WHO3, WHO4 and WHO5-6 COVID19 patients. For all boxplots the whiskers extend from the hinge to the largest and smaller values no further than 1.5 x interquartile range.

Neutrophils derived from WHO5-6 patients also had abnormalities in granule proteins notably a reduction in LTF (lactoferrin) and CAMP (cathelicidin; Fig. 3f), two of the three most abundant specific (secondary) granule proteins. This translated into a significant reduction in the protein content of specific granules in WHO4 and WHO5-6 patients compared to controls (Fig. 3g). Granule production is a hierarchical process linked to maturity, with azurophilic granules being the first granules produced follow by specific and tertiary granules and secretory vesicles (SV). Our data showed no significant changes in the protein content of azurophilic granules, ficolin granules, gelatinase granules or secretory vesicles (S.Fig. 3), suggesting no direct link to maturity, but a potential link to degranulation. We did however see altered expression of some azurophilic granule proteins linked with immunomodulation namely a reduction in LYZ (lysozyme) (Fig. 3h) and an increase in the ARG1 (arginase 1) (Fig. 3i). These data highlight how SARS-CoV-2 infection results in neutrophils with distinct potential immunoregulatory capacities and distinct capacities to respond to cues from other immune cells, pathogens or cytokines.

### Transient interferon and altered metabolic signatures in neutrophils from SARS-CoV-2-infected individuals

To gain insight into changes in neutrophil proteomes related to disease progression, we examined neutrophils from COVID19 patients 7 days after recruitment into the study. At this timepoint there were still major differences between the proteomes from COVID19 patients versus controls, with 2,081 proteins significantly changed in abundance (S.Table 8). As with the day 1 samples, the magnitude of the changes in protein signatures in the day 7 samples associate with disease severity: 239 proteins changed in WHO3 (S.Table 9) patients compared to controls, 1,373 in WHO4 (S.Table 10) and 828 proteins changed in WHO5-6 patients (Fig. 4a, S.Table 11).

**Figure 4.**
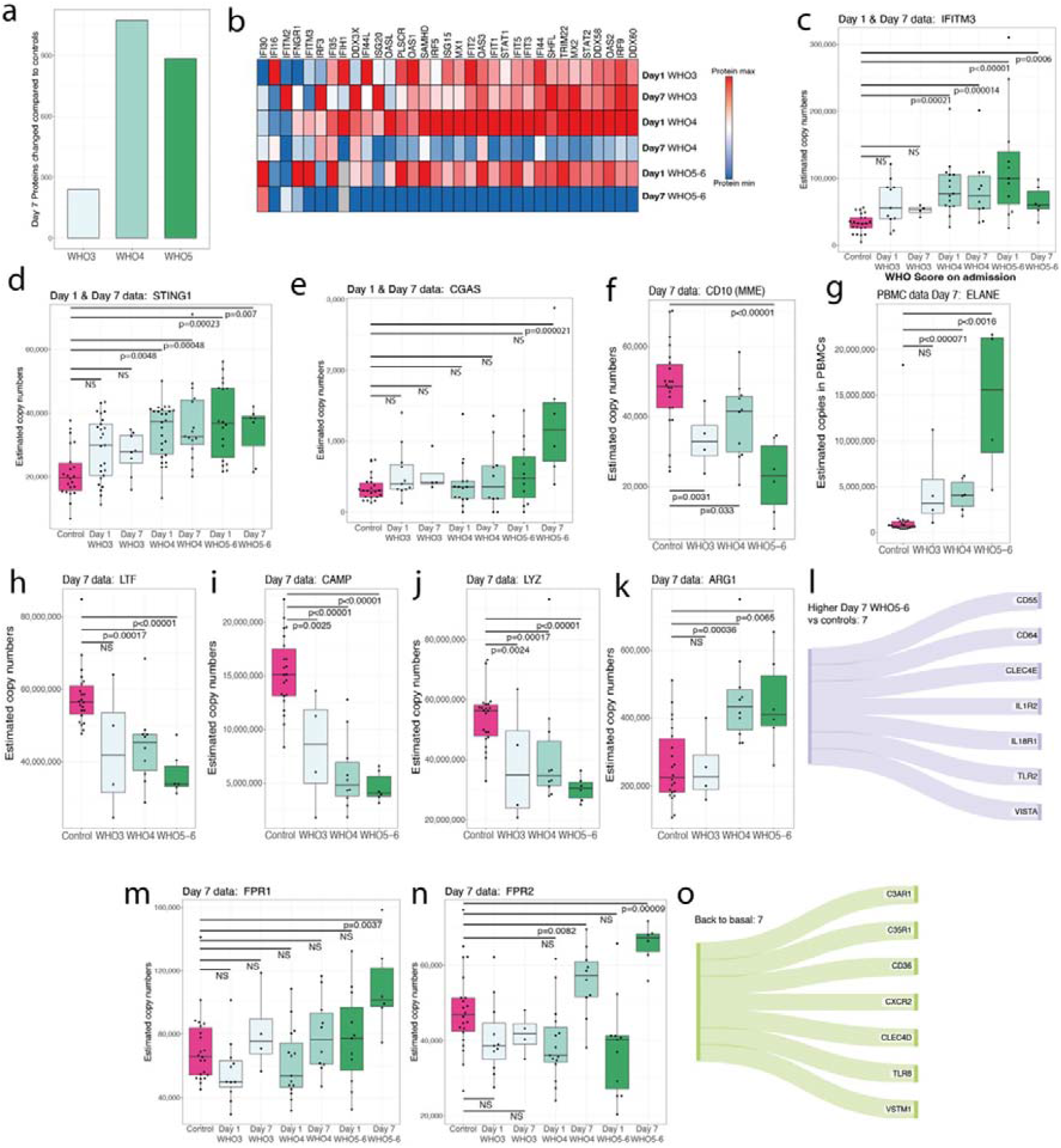
Transient and prolonged effects in the day 7 proteome: **(a)** Number of proteins significantly changed at day 7 in the WHO3 (moderate), WHO4 (severe) and WHO5-6 (critically ill) proteomes compared to controls. **(b)** Interferon response proteins across WHO3, WHO4 WHO5-6 patients for both day 1 and day 7. The data is normalised to the maximum value of each protein. Estimated protein copy numbers for **(c)** IFITM3, **(d)** STING1 and **(e)** CGAS across controls and WHO3, WHO4 and WHO5-6 patients in both day 1 and day 7. **(f)** Estimated protein copy numbers for CD10 across control, WHO3, WHO4 and WHO5-6 patients at day 7. **(g)** Estimated protein copy numbers in the PBMC proteomes for ELANE across control, WHO3, WHO4 and WHO5-6 patients at day 7. Estimated protein copy numbers for **(h)** LTF, **(i)** CAMP, **(j)** LYZ and **(k)** ARG1 across controls, WHO3, WHO4 and WHO5-6 patients at day 7. **(l)** Sankey diagrams showing immunomodulatory receptors which are significantly higher in the WHO5-6 patients at day 7 compared to controls. Estimated protein copy numbers for **(m)** FPR1 and **(n)** FPR2 across controls and WHO3, WHO4 and WHO5-6 patients in both day 1 and day 7. **(o)** Sankey diagrams showing immunomodulatory receptors which returned to basal levels (i.e. control levels) in the WHO5-6 patients at day 7 compared to controls. For all boxplots the whiskers extend from the hinge to the largest and smaller values no further than 1.5 x interquartile range.

A robust IFN-I response was still evident in WHO3 patients at day 7, showing no difference in the magnitude of the interferon signature compared to day 1 (Fig 4b). In contrast, the proteomes of WHO4 and WHO5-6 patients showed a prominent reduction of the IFN-I signature at day 7 (Fig. 4b). Of the interferon induced proteins elevated in the WHO5-6 group only IFITM3 (Fig. 4c), IFNGR1, STING (Fig. 4d) and cGAS (Fig. 4e) remained elevated at day 7 compared with controls.

In contrast to the reduced interferon signature, neutrophils and PBMCs from the WHO4 and WHO5-6 patients still showed hallmarks of immature/low-density neutrophils (Fig. 4f&g), and their neutrophils demonstrated persistent differences in granule proteins, with LTF, CAMP and LYZ still significantly reduced and ARG1 significantly increased (Fig. 4h-k). Furthermore, multiple signalling receptors that had been identified as markers of disease severity at day 1 remained elevated in the day 7 analyses such as TLR2, VISTA, IL1R2 and IL18R1 (Fig. 4l). Some receptors were only increased at day 7, such as the formyl peptide receptors FPR1 (Fig. 4m) and FPR2 (Fig. 4n), while half of the receptors were no longer different to the control neutrophils (Fig. 4o), including migratory receptors like C5AR1 and CXCR2, which were significantly reduced at day 1 but not day 7.

### Neutrophil proteomic signatures of recovered versus non recovered COVID19 patients

To explore longer term effects of SARS-CoV-2 infection on neutrophils we focussed on the proteomic analysis of neutrophils from COVID-19 patients 29 days after enrolment. For these analyses, patients were stratified into those who were at home and fully recovered (R-WHO1) and patients at home with persistent symptoms and limitations or still hospitalized (R-WHO2-3). Neutrophils from R-WHO2-3 patients showed significant changes in 1,111 proteins compared to neutrophils from control population (S-table 12). In contrast, only 404 proteins were significantly different between neutrophils from R-WHO1 patients versus controls (S-table 13, Fig. 5a). There were 268 proteins that were changed in neutrophils from both R-WHO1 and R-WHO2-3 patients, when compared to controls (Fig. 5b). These included increased expression of the CSF receptor CSF2RA (Fig. 5c), reduced abundance of ITAM adaptor FcRγ (Fig. 5d), reduced abundance of PYGL (Fig. 5e) and the migratory receptor LTB4R (Fig. 5f).

**Figure 5.**
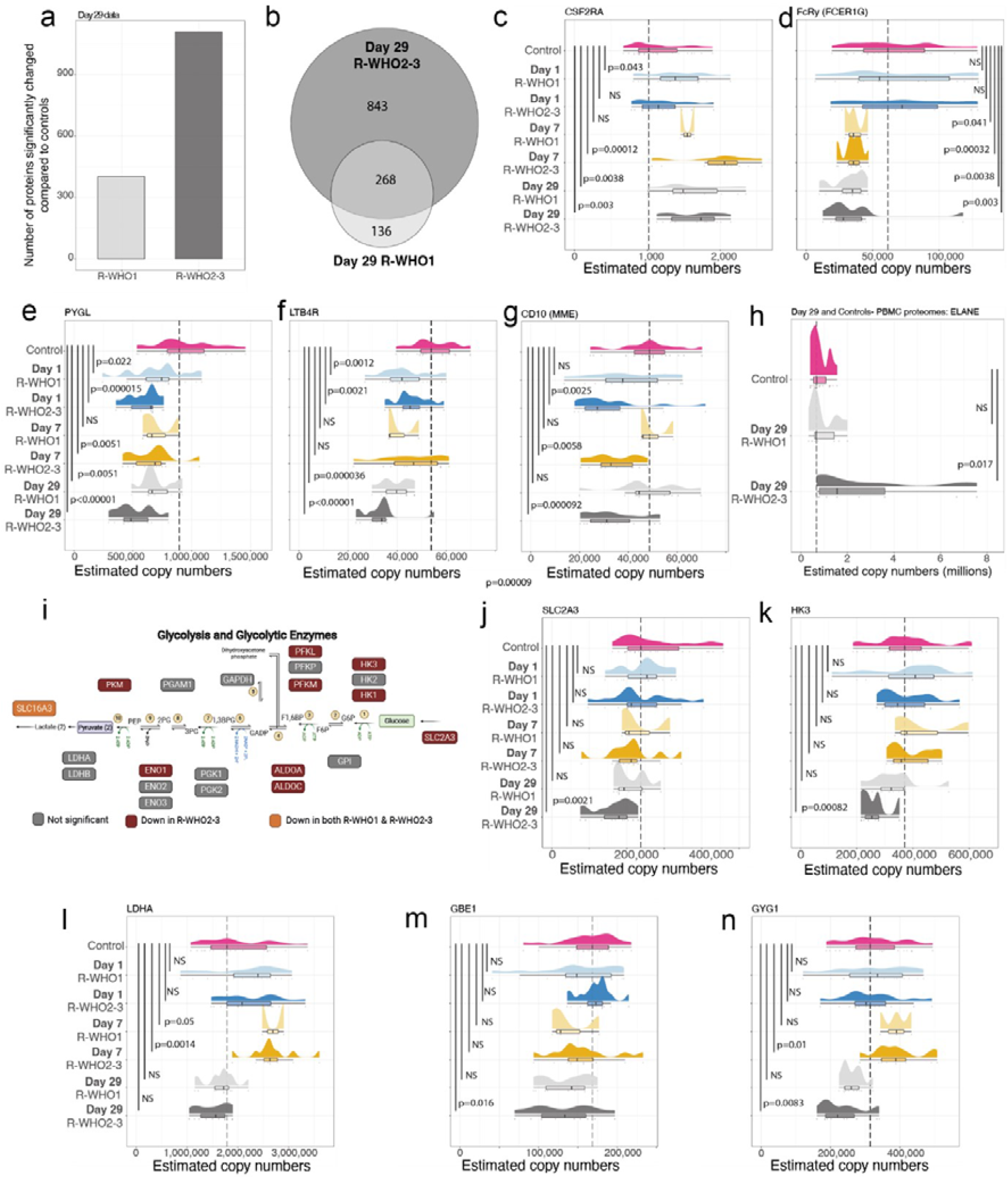
Metabolic changes in the neutrophil proteomes at day 29: **(a)** Number of proteins significantly changed in the neutrophil proteomes of R-WHO1 (recovered) or R-WHO2-3 (not recovered) patients at day 29 compared to controls. **(b)** Overlap of significantly changed proteins in the R-WHO1 and R-WHO2-3 day 29 proteomes. Rain cloud plot showing the estimated protein copy numbers for **(c)** CSF2RA, **(d)** FcRy (FCER1G), **(e)** PYGL, **(f)** LTB4R and **(g)** CD10 across controls, Day 1, Day 7 and Day 29 COVID19 patients stratified intp R-WHO1 and R-WHO2-3 (i.e. by day 29 WHO score). **(h)** Rain cloud plot showing the estimated protein copy numbers in the PBMC proteomes for ELANE across controls as well as Day 29 stratified in R-WHO1 and R-WHO2-3. **(i)** Glycolytic pathway highlighting proteins that were significantly reduced in abundance in the R-WHO2-3 patients. Rain cloud plot showing the estimated protein copy numbers for **(j)** SLC2A3, **(k)** HK3, **(l)** LDHA, **(m)** GBE1 and **(n)** GYG1 across controls, and Day 1, Day 7 and Day 29 COVID19 patients stratified into R-WHO1 and R-WHO2-3. All Raincloud plots include density plots as well as boxplots. For all boxplots the whiskers extend from the hinge to the largest and smaller values no further than 1.5 x interquartile range.

The data also revealed 843 proteins that were only changed in R-WHO2-3 neutrophils; these included the previously mentioned proteomic markers for immature neutrophils (Fig. 5g&h), with data from both the PBMC and neutrophil proteomes suggesting the presence of immature neutrophils was linked to delayed recovery. The PBMC proteome no longer indicated alterations in B cells in R-WHO2-3 patients, and no persisting differences in T cells in convalescent COVID19 patients as described in the literature^(14, 52, 53)^ (S.Fig. 4). Other characteristics of neutrophils from R-WHO2-3 patients related to changes in key metabolic proteins that control glycolysis, glycogenolysis and glycogen synthesis. These changes included reduced expression of rate limiting regulators of glycolysis like the glucose transporter SLC2A3, Hexokinase 3 and the lactate transporter SLC16A3 (Fig. 5i). Strikingly, the changes to glycolytic proteins in the day29 neutrophil populations were different to those seen in neutrophils at day1 and 7. For example, day1 and day7 neutrophils did not show decreased expression of SLC2A3 (Fig. 5j) or Hexokinase 3 (Fig. 5k). Additionally, there were glycolytic enzymes that were increased in expression in day7 neutrophils that were now at normal levels in the day29 neutrophils (eg lactate dehydrogenase) (Fig. 5l). One other difference in the metabolic proteome in the day29 neutrophils was reduced expression in R-WHO2-3 patients of enzymes that control glycogen synthesis such as the glycogen branching enzyme (GBE1; Fig. 5m) and glycogenin 1 (GYG1; Fig. 5n). Neutrophils from R-WHO2-3 patients displayed a metabolic proteome with reduced glycolysis and glycogenolysis, similar to what has previously been described in neutrophils from individuals with COPD, which associated with impaired neutrophil survival and antimicrobial capacity^(54)^.

One striking observation was that neutrophils from R-WHO2-3 patients which displayed a systematic reduction in the abundance of receptors that control neutrophil migration from the blood into sites of inflammation (Fig. 6a), including the sphingosine-1-phosphate receptor S1PR4 (Fig. 6b) and chemokine receptor CXCR2 (Fig. 6c). They also displayed reductions in the most abundant integrin subunits, CD18 (Fig. 6d) and CD11b (Fig. 6e), as well as CD11a and CD11c (S.Fig 5) along with SYK (Fig. 6f) a kinase that mediates integrin signalling. With the exception of CD11c, this reduced abundance was not present in neutrophil proteomes at day1 or 7 (Fig. 6d-f). One other insight was that the day29 neutrophil proteomes of R-WHO2-3 patients showed reduced abundance of inhibitory receptors of the C-Type-Lectin family and leukocyte immunoglobulin-like family (Fig. 6g). These receptors function to restrict signals that activate neutrophils^(55, 56)^. Moreover, R-WHO2-3 patients also displayed reduced expression of the SH-2-containing inositol 5’phosphatase, SHIP-1 (INPP5D; Fig. 6h) and the SH-2 containing tyrosine phosphatase SHP-1 (PTPN6; Fig. 6i), which also mediate inhibitory signals^(57, 58)^ and in which mutations have been linked to severe autoimmune disease^(59)^. Concurrently the activation marker CD64, was significantly higher in abundance in the proteomes of R-WHO2-3 patients compared to controls indicating persistent neutrophil activation (Fig. 6j). In the proteomes of neutrophils from R-WHO2-3 patients at day29 some of the changes observed at day1 and 7, like the increased abundance of ARG1, were no longer present (S.Fig. 6), however the reductions in specific granules (Fig. 6k) driven by LTF and CAMP persisted (S.Fig. 6). Furthermore, the proteomes of R-WHO2-3 patients displayed significant reductions in protein content of ficolin granules (Fig. 6l), a subset of highly mobilizable granules, distinct from the gelatinase granules^(60)^. As granule abundance is linked to maturity, we again considered whether these observed changes reflected increased prevalence of immature neutrophils in the R-WHO2-3 individuals. As previously mentioned, the last granules to be produced as neutrophils mature are the tertiary granules and secretory vesicles and these set of granules displayed no significant changes in protein content in the neutrophil proteomes from R-WHO2-3 patients (S.Fig. 7). The selective reductions in specific and ficolin granules would thus appear to be maturity independent.

**Figure 6.**
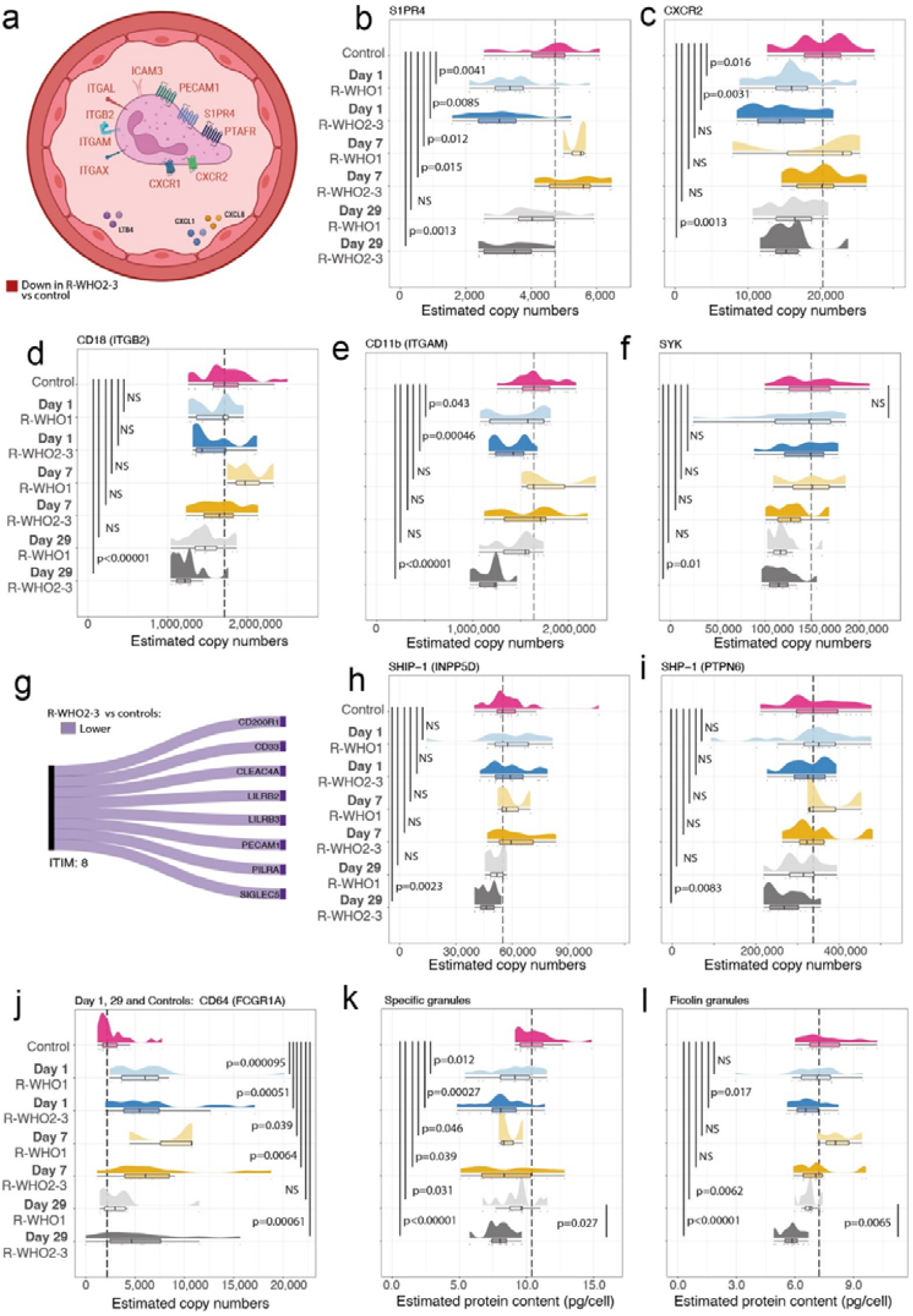
Neutrophil migratory and inhibitory machinery at day 29: **(a)** Migratory receptors and integrins that are exclusively reduced in abundance in the R-WHO2-3 patients compared to Controls. Rain cloud plot showing the estimated protein copy numbers for **(b)** S1PR4, **(c)** CXCR2, **(d)** CD18 (ITGB2), **(e)** CD11b (ITGAM) and **(f)** SYK across controls, Day 1, Day 7 and Day 29 COVID19 patients stratified into R-WHO1 (recovered at day 29) or R-WHO2-3 (not recovered at day 29). **(g)** Sankey diagram showing the inhibitory receptors that are significantly decreased in abundance in the neutrophil proteomes of R-WHO2-3 patients. Rain cloud plot showing the estimated protein copy numbers for **(h)** SHIP-1 (INPP5D), **(i)** SHP-1 (PTPN6) and **(j)** CD64 (FCGR1A) across controls, Day 1, Day 7 and Day 29 COVID19 patients stratified in R-WHO1 and R-WHO2-3. Estimated protein content for all proteins contained primarily within **(k)** Specific Granules and **(l)** Ficolin Granules across controls, Day 1, Day 7 and Day 29 COVID19 patients stratified in R-WHO1 and R-WHO2-3. All Raincloud plots include density plots as well as boxplots. For all boxplots the whiskers extend from the hinge to the largest and smaller values no further than 1.5 x interquartile range.

## Discussion

This study represents a large-scale and comprehensive proteomic characterisation of hundreds of neutrophil proteomes studied in both health and disease. It provides a valuable resource to understand the phenotypes of human blood neutrophils from healthy individuals in addition to an in-depth proteomic map of changes to neutrophils during acute COVID19 and the immediate recovery period. Prior studies of small cohorts with COVID19-associated ARDS identified a IFN-I proteomic signature in neutrophils^(32, 61)^. The present data show that this is a core COVID19 signature present in the majority of neutrophil proteomes derived from COVID19 patients, regardless of disease severity. The current data also show that this signature can be transient and that the kinetics are divergent and depend on disease severity. Hence in the neutrophils of COVID19 patients with moderate disease there was a sustained interferon protein signature whereas in patients with severe COVID19 this signature more rapidly returned to levels similar to controls. This divergence suggests the potential for stratified therapeutic interventions. MX1 has been previously described as a marker for IFN-I activity in multiple sclerosis^(62)^, and a lateral flow test has been developed^(63)^. This could potentially be used to stratify critically severe patients lacking an IFN-I response who could benefit from IFNβ treatment.

We identified novel neutrophil markers of disease severity including the pattern recognition receptor TLR2 and the inhibitory receptor VISTA. The association of high VISTA levels on neutrophils with COVID19 disease severity is intriguing as it has been suggested that this is a target to manage excessive innate immune activation^(64)^. The present data suggest that VISTA could be a target to reprogram neutrophil responses in severe disease caused by respiratory viruses. The current study also noted metabolic changes in neutrophils associated with severity and recovery status. Critically ill patients display changes in abundance of proteins across a wide array of metabolic pathways, some of which are explained by the increased proportion of immature neutrophils in patients with severe disease. There were also metabolic changes associated with disease severity (e.g. LDHA), which are linked to hypoxia^(44, 47)^ and NET production^(65)^, and we observed that glycogenolysis was decreased across all timepoints.

COVID19 causes prolonged illness in a subset of patients and a novel aspect of this study was the analysis of neutrophil proteomes during recovery. At day 29 post-enrolment, patients that had fully recovered from COVID19 displayed neutrophil proteomes increasingly similar to controls while patients who had not recovered by day 29 had abnormal metabolic profiles distinct from the changes seen in neutrophils isolated from patients on days 1 and 7. Neutrophils have been shown to depend on glycolysis for energy production when the environment is nutrient rich^(17, 66-68)^, and to default to glycogen breakdown when it is not^(54)^. These two metabolic pathways are vital to neutrophil functions, and our data show significantly lower abundance of key rate-limiting proteins of both glycolysis and glycogenolysis in the proteomes of non-recovered patients. Reductions in both pathways impair the bioenergetic capacity of neutrophils and have been shown to lead to impaired killing and impaired survival capacities in chronic disease^(54)^.

The dysfunctional phenotype of post-COVID19 neutrophils was not limited to metabolism. Peripheral blood neutrophils depend on signalling receptors and integrins to recognise migratory signals and perform the extravasation process. The neutrophil proteomes of non-recovered patients displayed a systematic reduction in migratory receptors, from the chemokine and complement receptors to spingosine-1-phosphate receptors. They also showed significantly reduced abundance of subunits of the Mac-1 and LFA-1 complexes that mediate leucocyte extravasation^(69)^. These changes suggest impaired capacity of neutrophils to migrate from the blood into the sites of inflammation, which may theoretically make patients post-COVID19 vulnerable to secondary infections; as mutations that affect function or abundance of CD18 in human neutrophils cause Leucocyte Adhesion Deficiency (LAD) and result in increased susceptibility to bacterial infections^(70)^. Non-recovered COVID19 patients also displayed a systematic reduction in the abundance of inhibitory receptors and phosphatases which are required to limit neutrophil activation^(55-58, 71)^. This reduction in the inhibitory machinery might explain the persistent degranulating phenotype present in peripheral blood neutrophils, which can have deleterious consequences as activated neutrophils in the bloodstream have been described as potentially lethal^(17, 72)^, however this programmed disarming of neutrophils could also be a mechanism to protect tissues^(73)^.

In conclusion, COVID19 remains a risk to global health, and we believe our data has identified a core neutrophil proteomic signature associated with acute disease and identified neutrophil receptors linked to disease severity which could be potential therapeutic targets. Furthermore, the investigation characterised a molecular phenotype linked to delayed recovery which potentially represents a mechanism relating to long covid and the extended symptoms COVID19 patients can experience.

## Data Availability

All the mass spectrometry files, as well as the processed search result files are available at PRIDE

## Data availability

All the mass spectrometry files, as well as the processed search result files are available at PRIDE^(74)^ under the identifier PXD0…

## Acknowledgements

We would like to thank Sarah Walmsley for insightful discussions relating to the data. We thank the team members of the proteomics facility at the University of Dundee, in particular Kenneth Beatie, Douglas Lamont and Abdelmadjid Atrih, for support in processing the mass spectrometry samples.

## Notes

Funding: This work was funded by the Chief Scientist Office Rapid Response COVID19 Research Grant (COV/DUND/20/01) and the UK Coronavirus Immunology Consortium (MR/V028448) and the Wellcome Trust (205023/Z/16/Z). DAC is supported by a Wellcome Trust Principal Research fellowship (097418/Z/11/Z). JDC is supported by the GSK/Asthma and Lung UK Chair of Respiratory Research (C17-2) and a Scottish Senior Fellowship from the Chief Scientist Office (SCAF17/03).

### Competing Interest Statement

James D. Chalmers has received research grants from AstraZeneca, Boehringer Ingelheim, GlaxoSmithKline, Gilead Sciences, Novartis and Insmed; and received consultancy or speaker fees from AstraZeneca, Boehringer Ingelheim, Chiesi, GlaxoSmithKline, Insmed, Janssen, Novartis and Zambon.

### Funding Statement

This work was funded by the Chief Scientist Office Rapid Response COVID19 Research Grant (COV/DUND/20/01) and the UK Coronavirus Immunology Consortium (MR/V028448) and the Wellcome Trust (205023/Z/16/Z). DAC is supported by a Wellcome Trust Principal Research fellowship (097418/Z/11/Z). JDC is supported by the GSK/Asthma and Lung UK Chair of Respiratory Research (C17-2) and a Scottish Senior Fellowship from the Chief Scientist Office (SCAF17/03).
 

### Author Declarations

The East of Scotland Research Ethics Committee gave ethical approval for this work (reference number 20/ES/0055)

