## Supplemental methods figure and tables for "Neutrophil proteomics identifies temporal changes and hallmarks of delayed recovery in COVID19"

**Affiliations**

***Methods:***

The PREDICT-COVID19 study was a prospective observational case-control study in a single UK NHS Hospital (Ninewells Hospital, Dundee). Patients with suspected or confirmed COVID19 were enrolled within 96 hours of hospital admission with SARS-CoV-2 infection confirmed by RT-PCR performed on combined oropharyngeal and nasopharyngeal swabs. Written informed consent was provided by all participants. The primary objective was to use proteomics to understand neutrophil function and heterogeneity in COVID19 as well as identifying biomarkers associated with outcomes. The secondary objective was to compare patients with SARS-CoV-2 infection to two control populations; patients presenting with acute respiratory tract infections not due to SARS-CoV-2 infection, and age and sex matched non-infected controls recruited without evidence of infection.

Research blood sampling was performed on day 1, and for confirmed SARS-CoV-2-positive participants additionally on days 4, 7, 15 and 29 while in hospital. Timing of blood sampling was standardised (between 0900 and 1100h each day). A subset of participants with COVID19 returned to the clinical research centre at Ninewells Hospital for sampling at day 28/29 while outpatients. Inclusion criteria for cases were: hospitalisation due to COVID19, positive or suspected diagnosis of COVID19, age >16 years. Exclusion criteria were inability to provide informed consent, or if venepuncture or participation was considered not in the best interests of the patient by the investigator. Inclusion criteria for controls were age >16 years, no known current infection, judged as clinically stable by the investigator and able to give informed consent. Exclusion criteria for controls were known or past SARS-CoV-2 infection in the past 3 months, known contact with a COVID19 positive case in the preceding 14 days, any current infection, and any contraindication to venepuncture or participation in the study as judged by the investigator.

**Clinical outcomes**

Key clinical outcomes against which proteomic data were to be compared were mortality, requirement for mechanical ventilation, requirement for non-invasive ventilation, clinical severity on a 7-point ordinal scale as well as clinical outcome at day 29 on a 7-point ordinal scale.

**Neutrophil isolation** 20ml of peripheral venous blood was drawn into 2x10ml EDTA(K2) vacutainers using a 21G butterfly needle for cell isolation. Within a maximum of 2h of venepuncture, neutrophils were isolated using EasySep™ Direct Human Neutrophil Isolation Kit (STEMCELL Technologies #19666) utilising negative immunomagnetic selection, as per the manufacturer's instructions.

In brief, 50µl Selection Cocktail and Direct RapidSpheres™ were added per ml of blood and incubated for 5 min. DPBS (without $Mg^{2+}/Ca^{2+}$) containing 1mM EDTA was added to a final volume of 50ml and the tube inserted into an Easy50 EasySep™ magnet (STEMCELL Technologies #18002). After incubation for 10 min, the neutrophil-rich upper layer was transferred to a new tube and the same volume of RapidSpheres™ as in the first step was added. Following 5 min incubation, the tube was inserted into the magnet for a 5 then 10 min incubation, transferring the neutrophil-rich suspension to a new tube each time. 10µl of the final neutrophil suspension was removed for cell counting using a disposable haemocytometer, sealed using 65µl Gene Frames (ThermoScientific #AB0577). The remaining cell suspension was centrifuged (300g, 6 min) to pellet neutrophils. Plasma supernatant was removed and discarded, and the pellet was washed by gentle resuspension in 10ml DPBS followed by further centrifugation (300g, 6 min).

**PBMC isolation**

10ml of venous blood drawn into EDTA-coated tubes as described above was diluted 1:1 with DPBS, then gently layered onto 15ml of Lymphoprep™ solution (STEMCELL #07811) in a 50ml SepMate™ column (STEMCELL #85450), and centrifuged for 20 min (1200*g*). Subsequently, the resulting layer of PBMCs was decanted and 10µl of the cell suspension was removed for cell quantitation as detailed above. The PBMC suspension was centrifuged at 300*g* for 8 mins, the cell pellet was then resuspended in 10 ml of DPBS as a washing step to remove contaminating free proteins, and centrifuged again (300*g*, 8 min).

**Cell sample storage**

The washed neutrophil or PBMC pellet was resuspended in DPBS to achieve a cell concentration of $5\times10^6$ cells/ml. 1ml of the suspension was transferred into an Eppendorf Protein LoBind tube and cells were pelleted (300 g, 5 min). The supernatant was discarded and cell pellet immediately frozen by inserting the tube into a pre-cooled metal block at -80°C. Pellets were stored at -80°C until lysis for liquid-chromatography mass spectrometry (LC-MS).

**Sample preparation for LC-MS**

To minimise batch effects of lysis buffers and conditions, stored cell pellets ($5\times10^6$ cells) were lysed in batches of 50–100 samples. For this, 400µl of freshly-prepared lysis buffer was added (5% sodium dodecyl sulphate (SDS), 10mM tris(2-carboxyethyl) phosphine hydrochloride (TCEP) and 50mM triethylammonium bicarbonate (TEAB; pH8.5) in dH2O) at RT. Samples were vortexed at 20,000 rpm for 10s, twice, at 5 min intervals. Samples were shaken at 500 rpm at room temperature for 5 minutes and then boiled at 98 C for 5 minutes. Samples were allowed to cool before sonicating for 30 cycles (30 seconds on and 30 seconds off). 1 µl of benzonase (Millipore) was added to each sample and samples incubated at 37 C for 15 minutes. Samples were alkylated with the addition of iodoacetamide to a final concentration of 20 mM and incubated for 1 hour in the dark. During the alkylation step, protein was quantified using the EZQ assay (Thermo Fisher). 100 µg of protein was loaded onto S-Trap mini columns[1] (Protifi) and processed following the manufacturer's instructions. Digests were performed using 5 µg trypsin/sample at 47 C for 2 hours. Peptides were eluted from S-Trap columns firstly with 50mM ammonium bicarbonate, followed by 0.2% aqueous formic acid and lastly with 50% aqueous acetonitrile containing 0.2% formic acid. Eluted peptides were dried and suspended in 1 % formic acid and quantified using the CBQCA assay (Thermo Fisher).

**Liquid chromatography mass spectrometry (LC-MS) analysis**

1.5 µg of peptide for each sample was analysed on a Q-Exactive-HF-X (Thermo Scientific) mass spectrometer coupled with a Dionex Ultimate 3000 RS (Thermo Scientific) as described previously[2]. The following LC buffers were used: buffer A (0.1% formic acid in Milli-Q water (v/v)) and buffer B (80% acetonitrile and 0.1% formic acid in Milli-Q water (v/v)). 1 µg aliquot of each sample was loaded at 15 µL/min onto a trap column (100 µm × 2 cm, PepMap nanoViper C18 column, 5 µm, 100 Å, Thermo Scientific) equilibrated in 0.1% trifluoroacetic acid (TFA). The trap column was washed for 3 min at the same flow rate with 0.1% TFA then switched in-line with a Thermo Scientific, resolving C18 column (75 µm × 50 cm, PepMap RSLC C18 column, 2 µm, 100 Å). Peptides were eluted from the column at a constant flow rate of 300 nl/min with a linear gradient from 3% buffer B to 6% buffer B in 5 min, then from 6% buffer B to 35% buffer B in 115 min, and finally to 80% buffer B within 7 min. The column was then washed with 80% buffer B for 4 min and re-equilibrated in 3% buffer B for 15 min. Two blanks were run between each sample to reduce carry-over. The column was kept at a constant temperature of $50^{o}$ C at all times.

The data was acquired using an easy spray source operated in positive mode with spray voltage at 1.9 kV, the capillary temperature at 250° C and the funnel RF at 60° C. The MS was operated in DIA mode using parameters previously described (Muntel et al, 2019) with some modifications. A scan cycle comprised a full MS scan (m/z range from 350-1650, with a maximum ion injection time of 20 ms, a resolution of 120 000 and automatic gain control (AGC) value of 5 × $10^6$). MS survey scan was followed by MS/MS DIA scan events using the following parameters: default charge state of 3, resolution 30.000, maximum ion injection time 55 ms, AGC 3 x $10^6$, stepped normalized collision energy 25.5, 27 and 30, fixed first mass 200 m/z. The inclusion list (DIA windows) and windows widths are shown below. Data for both MS and MS/MS scans were acquired in profile mode. Mass accuracy was checked before the start of samples analysis.

**DIA isolation windows**

| **Window** | m/z | Isolation window | Window | m/z | Isolation window |
|---|---|---|---|---|---|
| **1** | 383.375 | 66.8 | 24 | 670.5 | 14.5 |
| **2** | 423 | 13.5 | 25 | 684 | 13.5 |
| **3** | 435 | 11.5 | 26 | 697 | 13.5 |
| **4** | 446.5 | 12.5 | 27 | 710.5 | 14.5 |
| **5** | 458 | 11.5 | 28 | 725.5 | 16.5 |
| **6** | 469 | 11.5 | 29 | 741 | 15.5 |
| **7** | 480 | 11.5 | 30 | 756.5 | 16.5 |
| **8** | 490.5 | 10.5 | 31 | 773.5 | 18.5 |
| **9** | 501 | 11.5 | 32 | 791 | 17.5 |
| **10** | 512 | 11.5 | 33 | 808.5 | 18.5 |
| **11** | 523 | 11.5 | 34 | 827 | 19.5 |
| **12** | 533.5 | 10.5 | 35 | 846.5 | 20.5 |
| **13** | 544 | 11.5 | 36 | 866.5 | 20.5 |
| **14** | 554.5 | 10.5 | 37 | 887.5 | 22.5 |
| **15** | 565 | 11.5 | 38 | 910.5 | 24.5 |
| **16** | 575.5 | 10.5 | 39 | 935.5 | 26.5 |
| **17** | 586 | 11.5 | 40 | 962.5 | 28.5 |
| **18** | 597.5 | 12.5 | 41 | 992 | 31.5 |
| **19** | 609.5 | 12.5 | 42 | 1025 | 35.5 |
| **20** | 621.5 | 12.5 | 43 | 1063 | 41.5 |
| **21** | 633 | 11.5 | 44 | 1108.5 | 50.5 |

| | | | | | |
|---|---|---|---|---|---|
| **22** | 645 | 13.5 | 45 | 1391.625 | 516.8 |
| **23** | 657.5 | 12.5 | | | |

**Spectronaut 14 processing**

The raw DIA data was processed using Spectronaut[3] 14. As the whole dataset was too large to analyse in a single cell batch, the data was processed in multiple batches. Firstly, a DIA library was generated from 106 DIA raw files using Pulsar, this library is included in the PRIDE submisison. Protein inference was performed using Trypsin/P as Digest Rule, and Specific as Digest Type. PSM, Peptide and Protein FDR were set to 0.01. The library consisted of 126,065 precursors, 70,531 Peptides and 5,901 Protein Groups.

The data was subsequently searched against this library with the parameters personalised as follows: the decoy method was set to 'Inverse', the Precursor and Protein q-value Cutoff was set to 0.01, major grouping was set to 'Protein Group Id' and minor grouping to 'Stripped Sequence'. The major group quantity was set to 'Sum peptide quantity' and the minor group quantity to 'Sum precursor quantity'. The Major Group Top N and Minor Group Top N were unselected. The quantity MS-Level was set to 'MS2' and the quantity type set to 'Area', data filtering was set to 'Qvalue'. Cross Run Normalization was disabled as was the imputation strategy. The data search was split into 6 batches of less than 75 raw files, and the results were then combine using the 'SNE combine' feature. The individual SNE files and the spectral library are included in the PRIDE[4] submission under identified PXD

**Protein copy numbers**

Protein copy numbers were calculated in R based on the proteomic ruler[5]. The copy numbers for all patients are included in Supplemental Table 14.

**Protein content**

The estimated protein content was calculated based on the protein copy numbers and converted into picograms.

**Protein filtering**

Proteins identified by a single razor peptide were not included in the analysis. Furthermore, proteins were also filtered using the contaminant list provided by MaxQuant[6]. For the differential expression analysis, proteins needed to be detected in 2 samples or more.

**Statistical methods expanded**

The differential expression analyses were performed in R (v. 4.0.3) and the global p-values and fold changes were calculated via the Bioconductor package Limma[7] (v 3.46.0). The estimated copy numbers were $\log_2$ converted before being fed to the linear model function lmFit() which used the method='robust' parameter. The linear model was the evaluated using the empirical Bayes statistics for differential expression function eBayes() with the parameter set to robust= TRUE. The list of p-values generated were then fed into the Bioconductor package qvalue (v 2.22.0), which was used with the default parameters to produce the list of q-values.

**Significance levels**

Results were considered significant when their q-value≤0.05 in the global analyses. For aggregated protein categories, like specific granules, and for the 8 individual PBMC based proteins, results were considered significant with a p-value≤0.05.

#### Data batches

Neutrophil samples collected in the study were lysed for proteomic analyses in 4 main batches, therefore statistical analyses were performed in the interests of avoiding bias from batched processing. Batches 1, 2 and 4 contain data from control, LRTI and COVID19 patients. Batch 3 contained data derived exclusively from COVID19 patients. Comparisons of day 1 COVID19 and LRTI patients to controls used data from batches 1,2 and 4. Comparisons done between different day 1 COVID19 patients stratified by WHO score were done with batches 1,2,3 and 4. All analysis done on longitudinal data (day 7 and day 29) compared to controls was done on samples from batch 4. Finally the comparison of Day1 vs Day7 for COVID19 patients used batches 3 and 4.

#### Overrepresentation analysis

Overrepresentation analyses (ORA) were performed using webgestalt[8]. The background for all analyses was defined as all the proteins that were detected within the dataset. Only proteins that were detected with a q-value<0.05 were used for the ORAs. The analyses included 'Biological Process' and 'Cellular Component' as the gene ontology functional databases, and 'CORUM' as the network database. A minimum number of 5 genes per category was selected and the significant level was changed from Top 10 to using FDR<0.05

#### Figure generation

The boxplots and raincloud plots were all generated in R using ggplot2 (v 3.3.5), ggdist (v. 3.1.1), ggbeeswarm (v 0.6.0). Sankey diagrams were generated using sankeymatic (https://sankeymatic.com/build/ ), while the schematic diagrams were generated using biorender (https://biorender.com/ ). The heatmaps for this manuscript were generated using Morpheus (https://software.broadinstitute.org/morpheus/ ). The input was based on the median copy numbers and for each protein the data was normalised based on the maximum copy number displayed, leading to a scale from 0 to 1.

#### Granule proteins

Proteins were classified into the different granule subsets based on the work previously described by multiple proteomics experiments[9,10]. The full list of proteins is available at Supplemental Table 15.

#### MHC class II proteins

Boxplots showing the estimated copy numbers for all MHC-II proteins include are based on following proteins; HLA-DMB, HLA-DOA, HLA-DOB, HLA-DPA1, HLA-DRA, HLA-DRB1, HLA-DRB3 and HLA-DRB5.

### ***Supplemental Figures:***

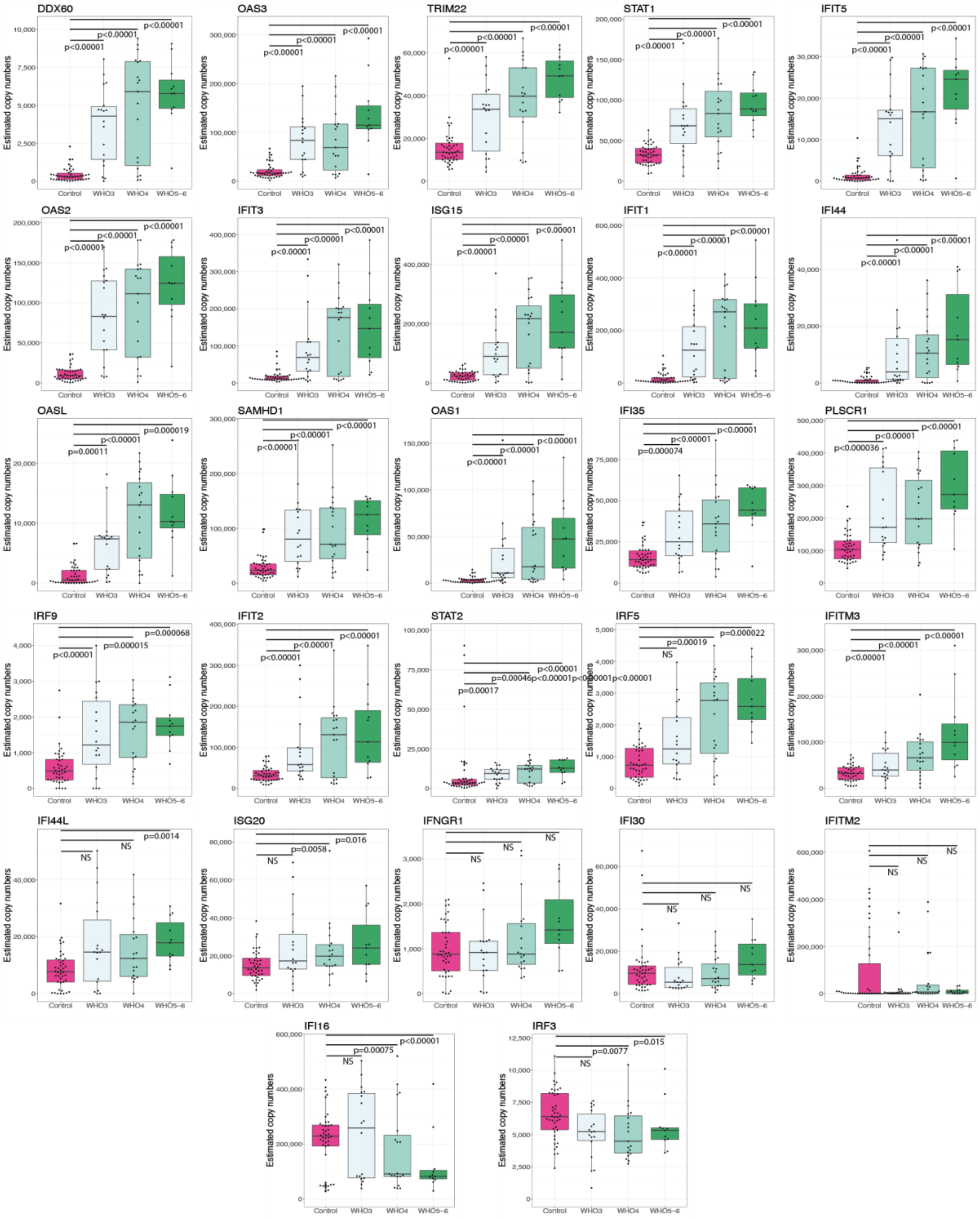


**Supplemental Figure 1:** Estimated copy numbers for the interferon response proteins at day 1 in the neutrophil proteomes across control participants and WHO3 (moderate), WHO4 (severe) and WHO5-6 (critically ill) COVID19 patients.

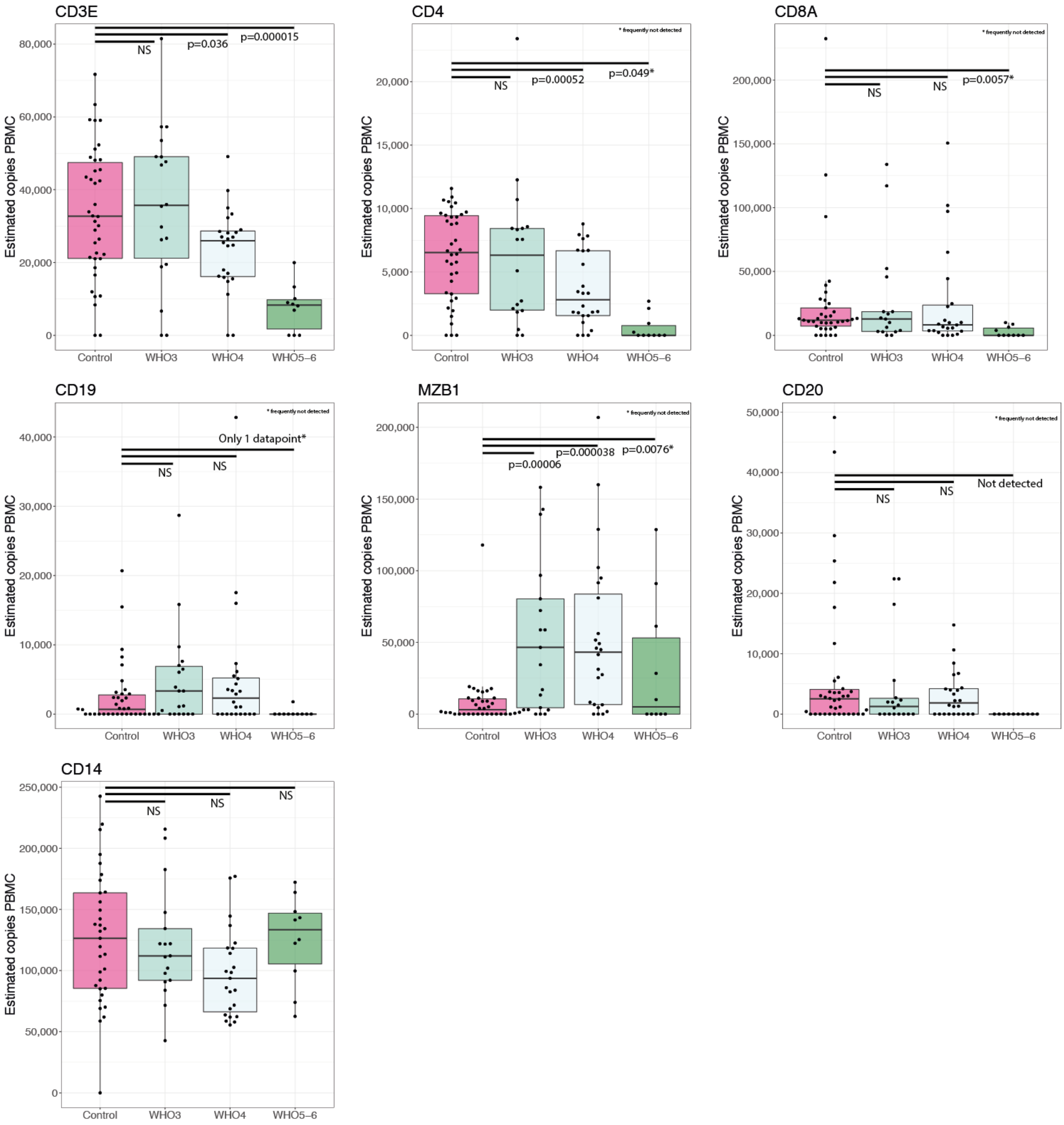


**Supplemental Figure 2:** Key cell type markers in PBMCs at day 1 derived from the PBMC proteomes across Control, WHO3 (moderate), WHO4 (severe) and WHO5-6 (critically ill) patients.

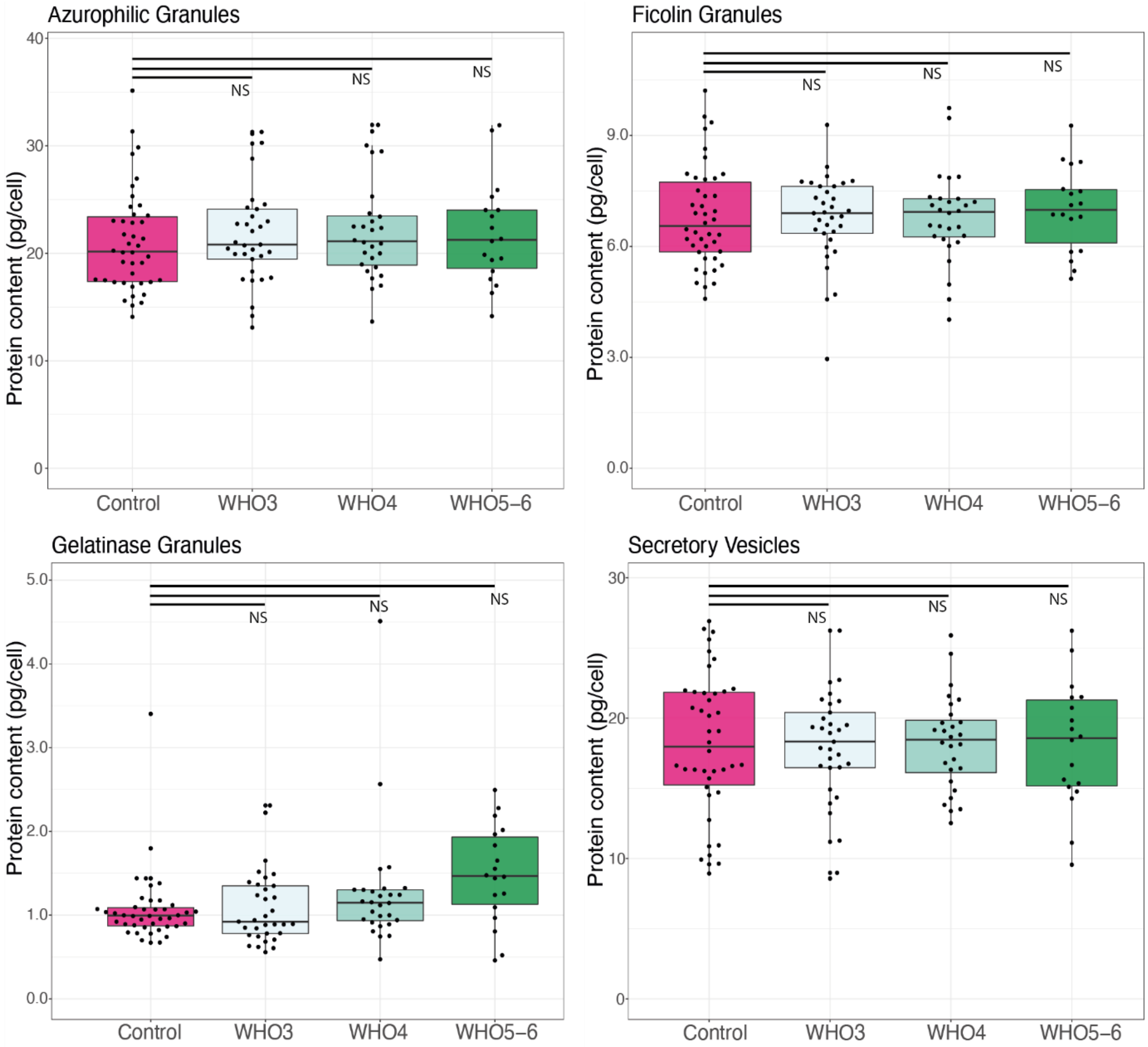


**Supplemental Figure 3:** Azurophilic (primary), ficolin and gelatinase (tertiary) granules and secretory vesicles at day 1 in neutrophil proteomes across control, WHO3 (moderate), WHO4 (severe) and WHO5-6 (critically ill) patients.

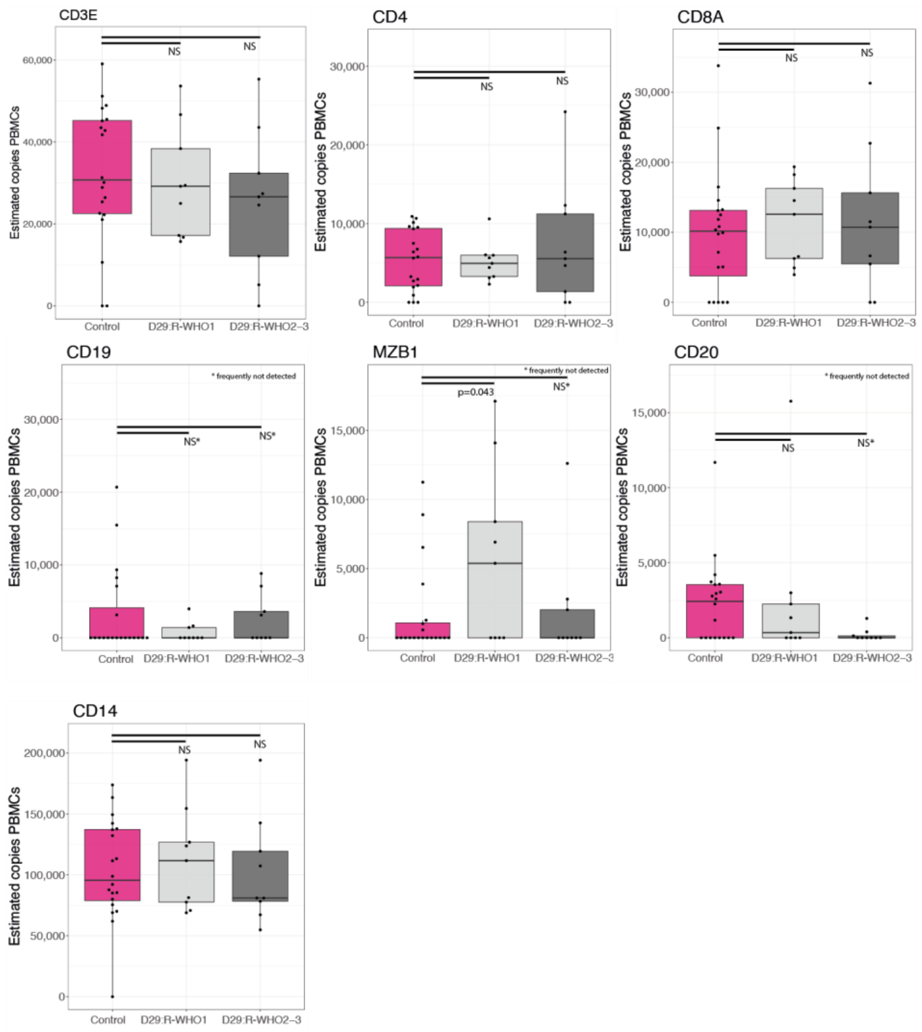


**Supplemental Figure 4:** Key cell type markers derived from PBMC proteomes across control participants, Day 29 R-WHO1 (recovered) and R-WHO2-3 (not recovered) COVID19 patients.

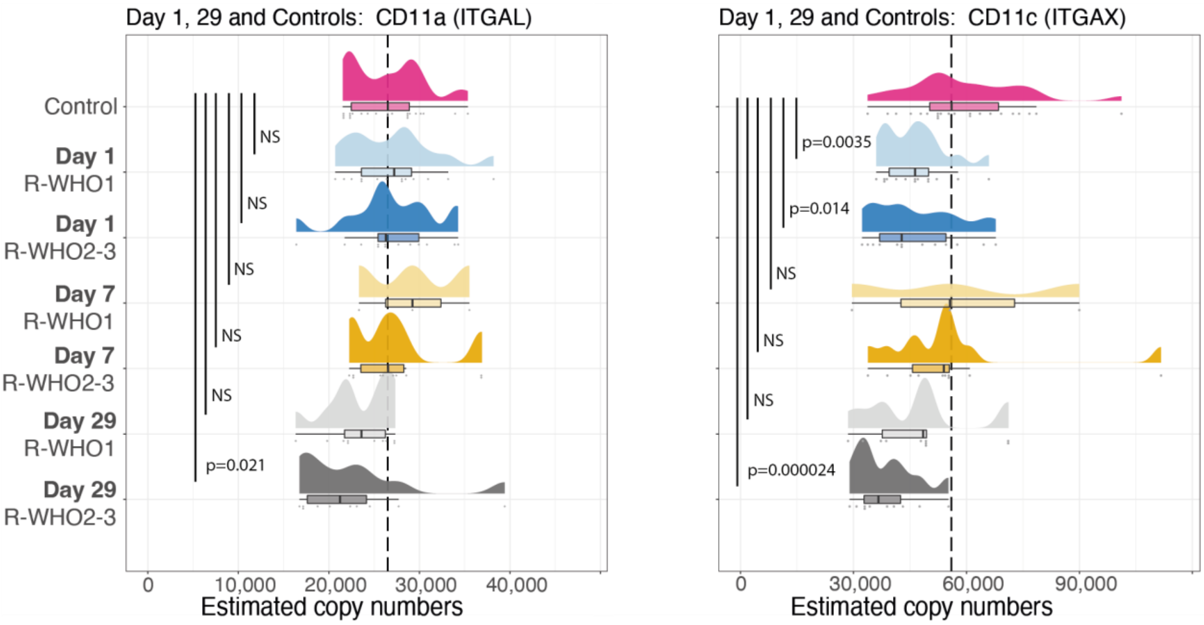


**Supplemental Figure 5:** Estimated copy numbers of CD11a & CD11c in neutrophil proteomes across control participants, Day 1 COVID19, Day 7 COVID19 and Day29 all stratified by R-WHO1 (recovered) and R-WHO2-3 (not recovered) patients.

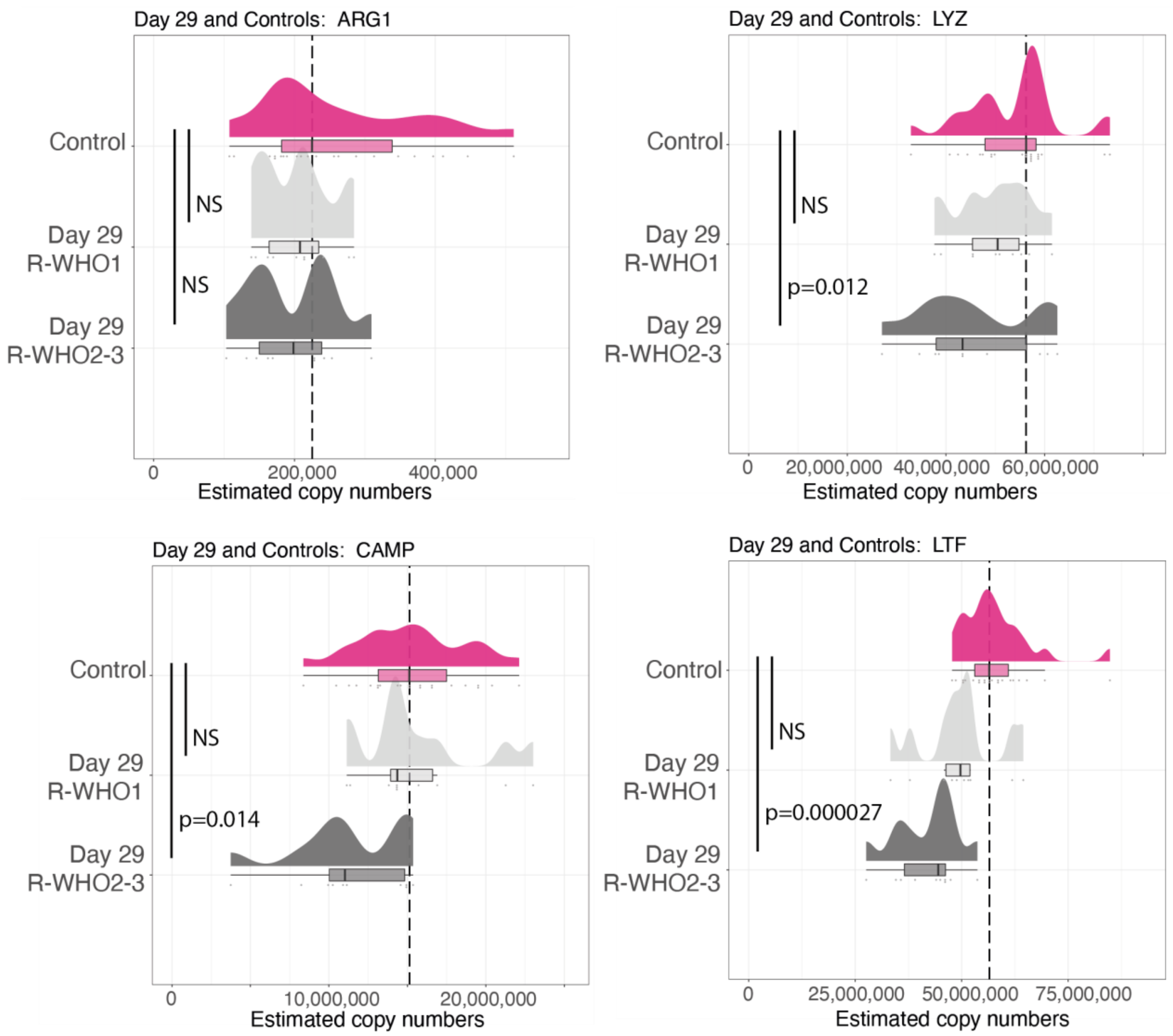


**Supplemental Figure 6:** Estimated copy numbers of LYZ, ARG1, CAMP & LTF at day 29 in the neutrophil proteomes of control group participants, and day 29 R-WHO1 (recovered) and R-WHO2-3 (not recovered) COVID19 patients.

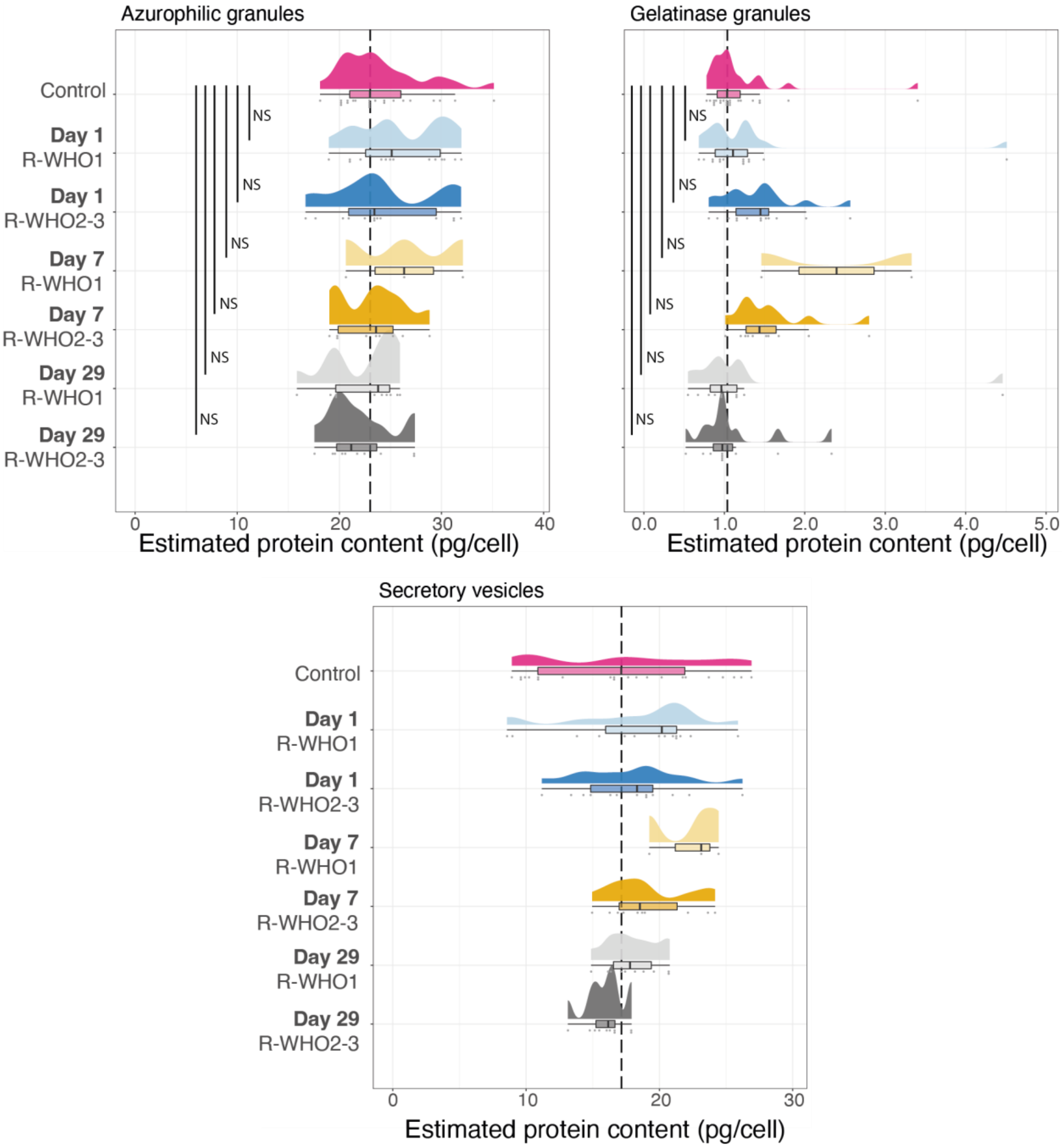


**Supplemental Figure 7:** Estimated protein content of azurophilic and gelatinase granules and secretory vesicles in the neutrophil proteomes of control participants at baseline, and Day 1 COVID19, Day 7 COVID19 and Day29 all stratified by R-WHO1 (recovered at day 29) and R-WHO2-3 (not recovered at day 29) patients.
